# A machine learning-based prediction of tau load and distribution in Alzheimer’s disease using plasma, MRI and clinical variables

**DOI:** 10.1101/2024.05.31.24308264

**Authors:** Linda Karlsson, Jacob Vogel, Ida Arvidsson, Kalle Åström, Olof Strandberg, Jakob Seidlitz, Richard A. I. Bethlehem, Erik Stomrud, Rik Ossenkoppele, Nicholas J. Ashton, Henrik Zetterberg, Kaj Blennow, Sebastian Palmqvist, Ruben Smith, Shorena Janelidze, Renaud La Joie, Gil D. Rabinovici, Alexa Pichet Binette, Niklas Mattsson-Carlgren, Oskar Hansson

**Affiliations:** Clinical Memory Research Unit, Department of Clinical Sciences in Malmö, Lund University, Lund, Sweden; Department of Clinical Sciences, SciLifeLab, Lund University, Lund, Sweden; Centre for Mathematical Sciences, Lund University, Lund, Sweden; Penn/CHOP Lifespan Brain Institute, University of Pennsylvania, Philadelphia, PA 19104, USA; Department of Psychiatry, University of Pennsylvania, Philadelphia, PA, 19104 USA; Department of Child and Adolescent Psychiatry and Behavioral Science, The Children’s Hospital of Philadelphia, Philadelphia, PA, 19104 USA; Institute for Translational Medicine and Therapeutics, University of Pennsylvania, Philadelphia, PA, 19104 USA; University of Cambridge, Department of Psychology, Cambridge Biomedical Campus, Cambridge, CB2 3EB, UK; Memory Clinic, Skåne University Hospital, Malmö, Sweden; Alzheimer Center Amsterdam, Department of Neurology, Amsterdam Neuroscience, Amsterdam UMC, Amsterdam, Netherlands; Department of Psychiatry and Neurochemistry, Institute of Neuroscience and Physiology, the Sahlgrenska Academy, University of Gothenburg, Mölndal, Sweden; Institute of Psychiatry, Psychology and Neuroscience, Maurice Wohl Institute Clinical Neuroscience, King’s College London, London, UK; Clinical Neurochemistry Laboratory, Sahlgrenska University Hospital, Mölndal, Sweden; Department of Neurodegenerative Disease, UCL Institute of Neurology, Queen Square, London, UK; UK Dementia Research Institute at UCL, London, UK; Hong Kong Center for Neurodegenerative Diseases, Clear Water Bay, Hong Kong, China; Wisconsin Alzheimer’s Disease Research Center, University of Wisconsin School of Medicine and Public Health, University of Wisconsin-Madison, Madison, WI, USA; Paris Brain Institute, ICM, Pitié-Salpêtrière Hospital, Sorbonne University, Paris, France; Neurodegenerative Disorder Research Center, Division of Life Sciences and Medicine, and Department of Neurology, Institute on Aging and Brain Disorders, University of Science and Technology of China and First Affiliated Hospital of USTC, Hefei, P.R. China; Department of Neurology, Memory and Aging Center, Weill Institute for Neurosciences, University of California, San Francisco, San Francisco, CA, USA; Department of Radiology and Biomedical Imaging, University of California, San Francisco, San Francisco, CA, USA

## Abstract

Tau positron emission tomography (PET) is a reliable neuroimaging technique for assessing regional load of tau pathology in the brain, commonly used in Alzheimer’s disease (AD) research and clinical trials. However, its routine clinical use is limited by cost and accessibility barriers. Here we explore using machine learning (ML) models to predict clinically useful tau-PET composites from low-cost and non-invasive features, e.g., basic clinical variables, plasma biomarkers, and structural magnetic resonance imaging (MRI). Results demonstrated that models including plasma biomarkers yielded the most accurate predictions of tau-PET burden (best model: R-squared=0.66-0.68), with especially high contribution from plasma P-tau217. In contrast, MRI variables stood out as best predictors (best model: R-squared=0.28-0.42) of asymmetric tau load between the two hemispheres (an example of clinically relevant spatial information). The models showed high generalizability to external test cohorts with data collected at multiple sites. Based on these results, we also propose a proof-of-concept two-step classification workflow, demonstrating how the ML models can be translated to a clinical setting. This study uncovers current potential in predicting tau-PET information from scalable cost-effective variables, which could improve diagnosis and prognosis of AD.

## Introduction

Alzheimer’s disease (AD) is the most common neurodegenerative disease, with prevalence continuing to increase world-wide.^1^ It is characterized by pathological aggregation of amyloid(A)-β plaques and neurofibrillary tau tangles in the brain. AD pathology can be reliably detected and quantified using fluid or imaging biomarkers, e.g., plasma (blood-based), cerebrospinal fluid (CSF) or positron emission tomography (PET).^2,3^ These AD biomarkers vary in availability, cost, invasiveness and comprehensiveness, and the most demanding methods are not feasible to implement in larger populations, even if they may be the most reliable and informative ones. Amyloid- and tau-PET are the only methods to spatially resolve AD pathology in the living human brain but is expensive and not readily available in most clinical settings. Conversely, plasma biomarkers are or will likely be highly accessible and minimally invasive, and although capturing a variety of biological aspects related to AD, they lack detail on spatial resolution of the pathology. Structural magnetic resonance imaging (MRI) represents a relatively lower-cost alternative to spatially measure *in vivo* neurodegeneration, but is substantially less specific to AD pathology compared to PET.^4–7^ With emerging disease-modifying therapies for AD^8,9^, including tau-targeting therapies under development^10^, it is crucial to find ways to replace PET scans due to their relative inaccessibility and costliness, while still preserving the ability to estimate the load and distribution of AD pathology as accurately and comprehensively.^11,12^

In this work, we specifically aim to address the problem of limited global access to tau-PET. Tau-PET is often considered a state-of-the-art outcome in AD research studies, a very accurate prognostic tool in AD patients, and frequently used in clinical trials for AD during screening and to monitor treatment effects.^11,13–15^ Furthermore, tau-PET provides information beyond what can be captured by other imaging modalities or fluid biomarkers, including *in-vivo* assessment of Braak stages, which indicates the progression of tau pathology throughout the brain.^16,17^ Even though tau pathology often spreads in a distinct spatial pattern, recent work has provided insights into the heterogeneity of the disease by characterizing different subtypes of the tau deposition patterns, which can be of relevance for an individually adapted prognosis.^18,19^ Asymmetry in tau accumulation between the two hemispheres can for example offer clues about the lateralization of pathology within the brain.^20–22^ Such asymmetry can give rise to atypical clinical profiles, related to for example behavioral and cognitive differences,^23^ and has been shown to relate to faster disease progression.^18^

Here, we approach the problem of limited tau-PET accessibility with machine learning (ML) models, trained to predict tau-PET information from low-cost and non- or minimally invasive variables only. We aim to understand the scope of accessible features as tools predictive of tau pathology (load and distribution), and to explore the potential of their expanded utility in a context where invasive and/or expensive procedures are not available. Specifically, we investigated how well two different tau-PET composites can be predicted from basic clinical variables, plasma AD biomarkers and structural MRI-derived morphology. These two composites were: 1) tau load in the temporal cortex (an early, sensitive, and commonly used composite of tau deposition in AD) and 2) hemispheric asymmetry of temporal lobe tau load in tau positive participants (as an example of clinically relevant spatial tau PET information). Toward this purpose, we created a flexible and comprehensive ML pipeline, designed to handle both different feature types and different number of input features for classification and regression tasks. We evaluated the results on an unseen test set and multiple external cohorts. We further investigated feature contribution for increased model interpretability and transparency. Lastly, we created a two-step pipeline proposed to simulate how the findings could be implemented in a clinical setting without access to tau-PET.

## Results

We included 1195 participants from the BioFINDER-2 cohort (BF2), consisting of individuals with normal cognition (NC, n=321), subjective cognitive decline (SCD, n=153), mild cognitive impairment (MCI, n=236), dementia (n=232, of which 136 were AD and 96 non-AD cases), or another neurodegenerative disease (n=6). All participants had undergone tau-PET, and they were randomly split into an 80% training set (n=948) and 20% test set (n=247). For external validation, we included 147 participants with tau-PET scans from the BioFINDER-1 cohort (BF1), and specifically tau-PET *positive* participants from the ADNI cohort (n=136), the UCSF-ADRC cohort (n=144), the OASIS3 cohort (n=46) and the A4 cohort (n=45). Demographic information on all cohorts is provided in Supplementary Tab. 1. Note that the relevant plasma variables were not available to us in ADNI, UCSF-ADRC, OASIS3 and A4, preventing external validation for all models using plasma in these cohorts.

Two tau-PET indices were predicted: tau-PET Braak I-IV load, corresponding to a temporal meta-ROI, and tau-PET Braak I-IV laterality index. The Braak I-IV laterality index had values above 0 when higher tau load was present in the left hemisphere, and below 0 for higher tau load in the right hemisphere. The distributions of the outcome variables (in all cohorts) are provided in scatter plots in Fig. 1a-1b, together with examples of low/high load and symmetric/asymmetric tau-PET scans in Fig. 1c.

**Fig. 1:**
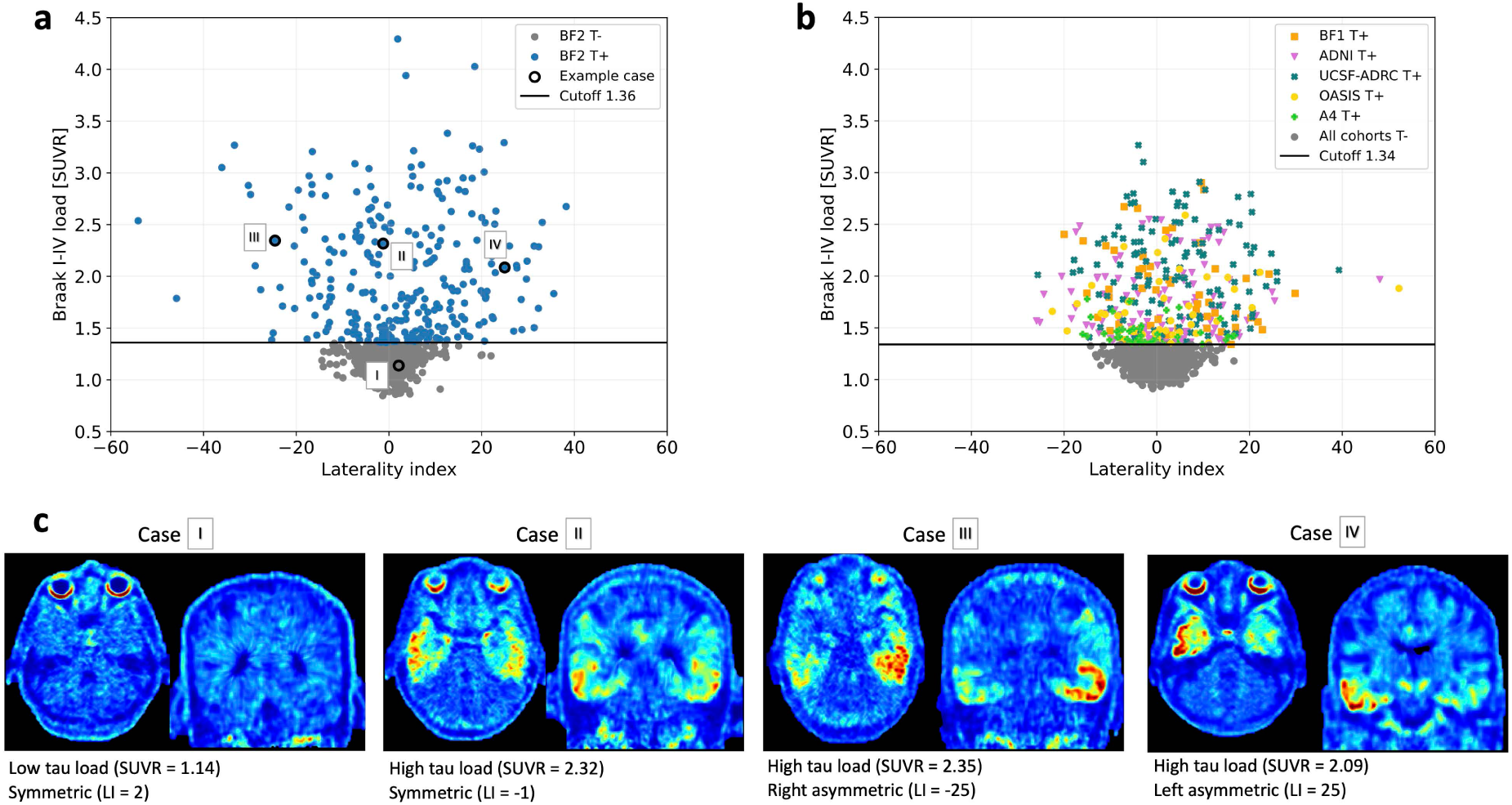
Distributions and example cases of the tau-PET composites to be predicted in machine learning models. Distribution of the two outcome variables tau-PET Braak I-IV load and laterality index (L>R) in a) BF2 and b) external test sets. Note that different tau-PET tracers were used in the cohorts ([^18^F]RO948 in BF2 and [18F]flortaucipir in the others), resulting in slightly different cutoffs for positivity (1.36 and 1.34). In c) tau-PET example cases of low/high tau load, and symmetric/asymmetric tau distribution from BF2 are provided (correspondingly marked out in a). *Abbreviations: LI (laterality index), SUVR (standardized uptake value ratio)*

### A flexible machine learning pipeline

To search for high-performing hyperparameter-tuned ML estimators we implemented a flexible ML pipeline, designed to handle different data types, different number of input features, and different evaluation metrics, for both classification and regression tasks. The pipeline tested with 10-fold cross-validation several feature selection steps (aimed to in a data-driven manner find appropriate dimensionality reductions robust to the varying number of input features and/or skewed proportions of certain types of input features) and ML estimators in a combined grid-search and Bayesian optimization set-up (Fig. 2 and Supplementary Tab. 2).

**Fig. 2:**
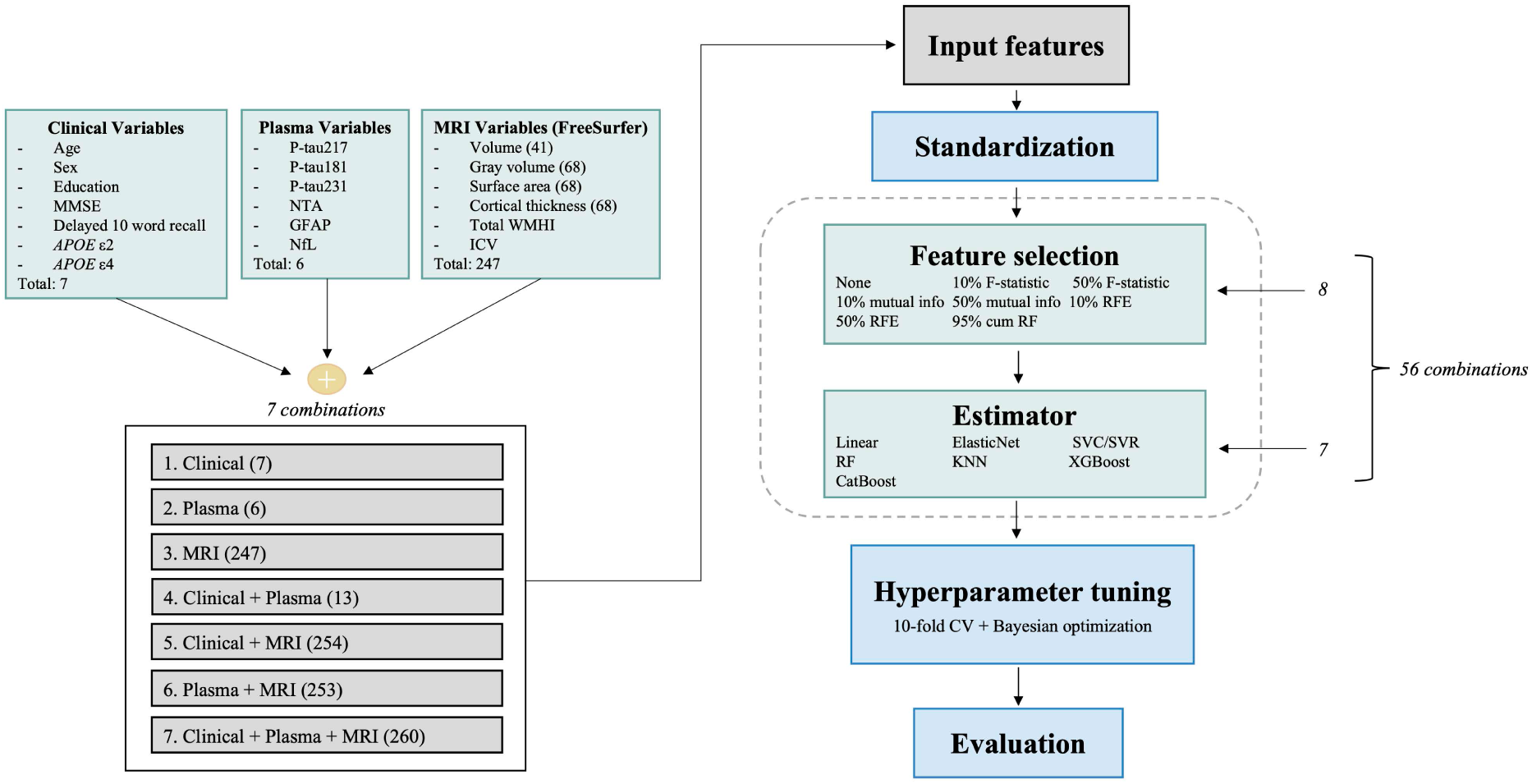
A rigorous and flexible machine learning pipeline created to compare combinations of input features in regression and classification tasks. Three different blocks of variables (clinical, plasma and MRI) were tested separately and in combination as input features in a pipeline created to search for high performing ML models for regression and classification tasks. The pipeline consisted of a combined grid search (feature selection and estimators, in total 56 combinations) and Bayesian optimization (hyperparameter tuning, in a 10-fold cross-validations setting) approach with the best score, corresponding tuned hyperparameters and computational time for all 56 combinations as output. The pipeline could handle different input and output variables, evaluation metrics and both regression and classification tasks, making it applicable to any such ML task. Code was written in Python and is available on https://github.com/DeMONLab-BioFINDER/karlsson-predict-taupet. *Abbreviations: RFE (recursive feature elimination), cum RF (cumulative random forest feature importance), SVC/SVR (support vector classifier/regressor), RF (random forest), KNN (K-nearest neighbor), CV (cross validation), WMHI (white matter hyperintensities), MMSE (mini mental state examination), ICV (intracranial volume)*

We evaluated several clinically accessible input features for each prediction task to search for the best performing combinations. The input features were divided a priori into three feature blocks: *Clinical Variables* (n_features_=7), *Plasma Variables* (n_features_=6), and *MRI Variables* (n_features_=247). Clinical variables were basic demographics (age, sex, education level, number of *APOE* ε2 alleles, and number of *APOE* ε4 alleles) as well as two cognitive tests (mini mental state examination MMSE and delayed 10-word recall). Plasma biomarkers were phosphorylated (P)-tau217, P-tau181, P-tau231, N-terminal containing tau fragments (NTA), glial fibrillary acidic protein (GFAP) and neurofilament light (NfL). Structural MRI features were derived from the FreeSurfer pipeline (v.6.0, http://surfer.nmr.mgh.harvard.edu/) including volumes, surface areas and thicknesses of cortical regions, volumes of subcortical brain regions, total white matter hyperintensities (WMHI) and total intracranial volume (ICV). An overview of the correlation between all input features is displayed as a hierarchically clustered heatmap in Supplementary Fig. 1, with numerical details in Supplementary information. The three input feature blocks *Clinical*, *Plasma* and *MRI Variables* were analyzed separately or together in all possible combinations (seven in total), Fig. 2. Hyperparameter-tuning and model-selection was performed using cross-validation on the BF2 training set unless specified otherwise. Final evaluation was performed on the hold-out test set. Evaluation of generalizability was performed on BF1, and when feasible (for models without plasma), also in ADNI, UCSF-ADRC, OASIS3 and A4.

### Plasma biomarkers were superior for predicting the load of temporal lobe tau tangle pathology

The first tau-PET composite we predicted was continuous Braak I-IV tau load (i.e., standardized uptake value ratio [SUVR] in the commonly used temporal meta-ROI). The *Plasma Variables* feature block consistently produced the lowest mean squared error (MSE), both alone and in combination with the other feature blocks (Fig. 3a). Tree-based ML models (random forest [RF] and boosting regressors) were superior to linear, elastic net and K-nearest neighbor (KNN) for most feature combinations. Details on MSE, selected hyperparameters and computational times for all 56 combinations in the machine learning pipeline and every input feature block can be seen in Supplementary information.

**Fig. 3:**
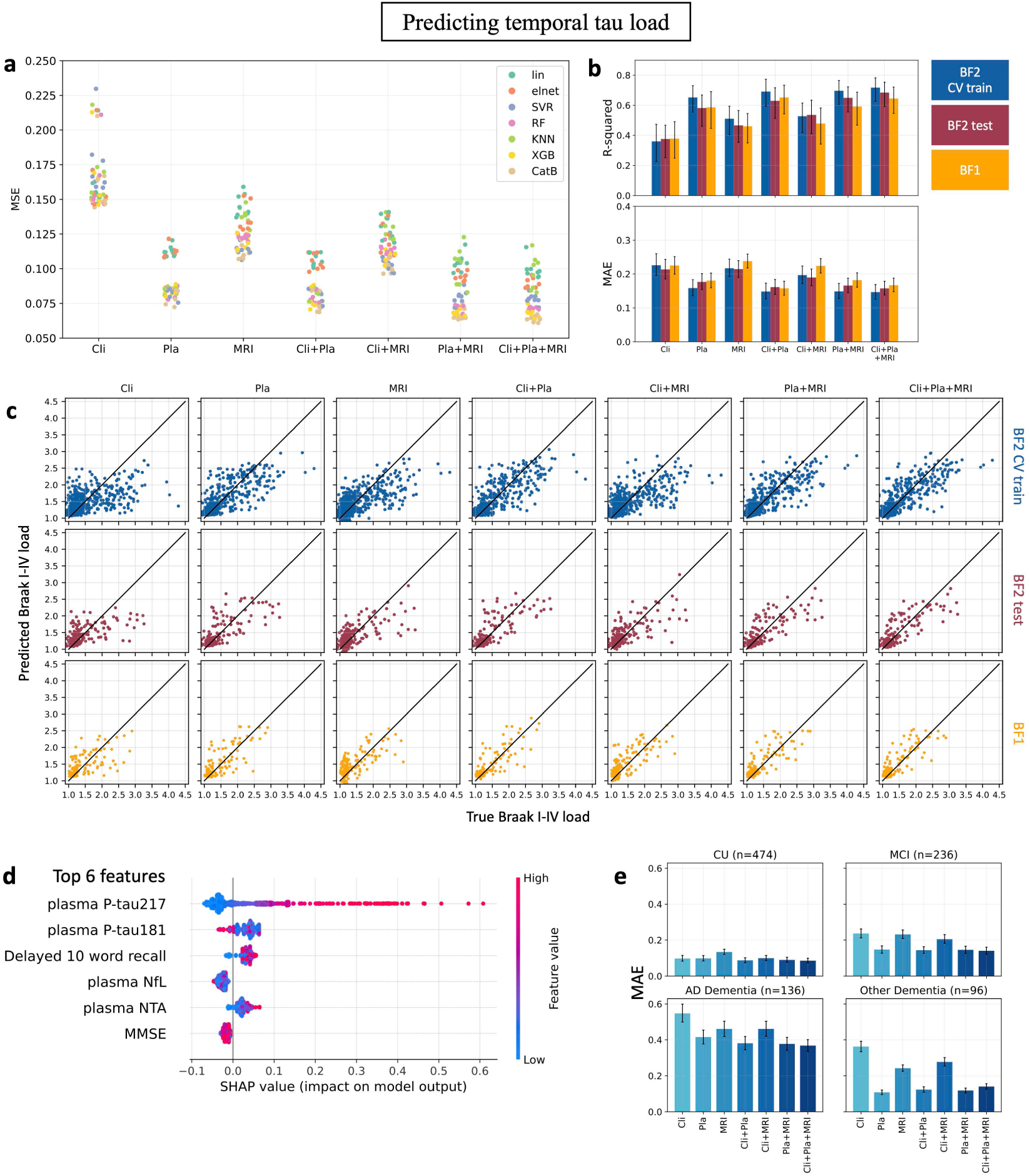
Plasma features were most predictive of temporal tau load. **a)** Mean squared error (MSE) of all 56 feature selection + estimator combinations (Fig. 2) for the different input feature blocks (clinical, plasma, MRI, and all combinations) when predicting temporal tau load in the cross-validated BF2 training set. Each scatter point is colored by estimator. The blocks that included plasma features consistently performed best, always in a CatBoost model. **b)** R-squared and mean absolute error (MAE) of the best pipeline combinations in a) for the cross-validated BF2 training set, BF2 test set and external test cohort BF1. **c)** Scatter plots of true vs predicted Braak I-IV (temporal tau) load for the best pipeline combinations in a) for the cross-validated BF2 training set, BF2 test set and BF1. **d)** top SHAP feature contributions in the model including all features, evaluated on the cross-validated BF2 training set. **e)** MAE performance of best pipeline combination in a) stratified by cognitive status in the cross-validated BF2 training set. *Abbreviations: Cli (Clinical), Pla (Plasma), MAE (mean absolute error), lin (linear regressor), elnet (elastic net regressor), SVR (support vector regressor), RF (random forest regressor), 2KNN (K-nearest neighbor regressor), XGB (XGBoost regressor), CatB (CatBoost regressor), CU (cognitively unimpaired), MCI (mild cognitive impairment), AD (Alzheimer’s disease)*

For each of the seven combinations of feature inputs, the best model (lowest MSE in Fig. 3a and Supplementary Tab. 3) were selected for further evaluation in the BF2 test set and external cohort BF1 (note that ADNI, UCSF, OASIS and A4 were excluded from this part due to absence of plasma variables). Performances on the BF2 test set and BF1 were similar to the corresponding cross-validated training set performance (Fig. 3b and Supplementary Tab. 4). The best model in the cross-validated training set was a boosting regressor (CatBoost model) that from clinical, plasma and MRI utilized a 95% cumulative RF feature selection (R-squared = 0.72 and MAE = 0.15 SUVR). On unseen test data, it resulted in R-squared of 0.66-0.68 and MAE of 0.16-0.17 SUVR, indicating high generalizability. However, other feature combinations that included plasma variables but not MRI and/or clinical variables reached similar or even higher performance on the test data (Fig. 3b and Supplementary Tab. 4), highlighting how fluid biomarkers were the key attribute to predict tau load. This was also visible in scatter plots of the true versus predicted results from the best model for each feature block combination (Fig. 3c). Using just clinical variables resulted in limited predicted value (R-squared = 0.38). Using just MRI variables resulted in some predictive value but models tended to underestimate tau load (R-squared = 0.46-0.47). Adding clinical or MRI information to the plasma biomarker block led to only marginal improvement.

As many models within each feature block performed very similar in the BF2 training set (Fig. 3a), we investigated if this small MSE difference affected the ability of the model to generalize to unseen data. We compared the best five models within each input block combination (Supplementary Tab. 4 and Supplementary Fig. 2). The top five models performed similarly within each feature block, but the distinction between input feature blocks remained such that inclusion of plasma biomarkers always resulted in the best predictive performance.

As the outcome variable was imbalanced between low and high load cases, we stratified the evaluation based on diagnostic groups. In this way, the evaluation metric would be more representative of the varying performance along the distribution of tau-PET load. Predicting tau-PET load was easiest for cognitively unimpaired (CU) and mild cognitively impaired (MCI) participants with no to moderate tau-PET load (lowest error in Fig. 3e). Plasma variables yielded superior results in all diagnostic groups but stood out the most by accurately identifying low tau load in the non-AD dementia group (Fig 3e). Clinical (e.g., cognitive tests and basic demographics) and MRI variables were less efficient when differentiating neurodegeneration due to tau pathology versus another pathology (larger errors for other dementias and MCI). In the AD dementia group, adding clinical and MRI variables to plasma improved performance slightly, but at the expense of inferior performance in other dementias. We saw no improved performance in predicting tau-PET load in Aβ-positive individuals when re-running the pipeline after excluding all Aβ-negative individuals, suggesting that the large proportion of participants with low tau load did not impair the models’ predictive ability of participants with high tau load.

SHAP analysis was used to interpret which features contributed most to the prediction. This analysis showed that plasma P-tau217 was clearly the most important predictor of temporal tau-PET load (Fig. 3d). Plasma P-tau217 contributed more to predictions when it had higher concentrations (higher feature value in the Fig. 3d). For further model transparency, the SHAP interaction plots for the features in Fig. 3d are provided in Supplementary Fig. 3.

Since plasma P-tau217 alone stood out as most informative during the prediction of tau-PET load, we compared the best ML models against a simple linear regression model using only P-tau217 as predictor. ML models without any plasma variables usually performed worse than the simple plasma P-tau217 linear model. Models combining the plasma feature block with clinical and/or MRI variables provided significantly better predictive performance than the simple model for the BF2 test set (ΔR2=0.13-0.18, ΔMAE=0.031-0.039 SUVR). In BF1, only the “fluid + clinical” model significantly outperformed the plasma P-tau217 model, and only regarding MAE (ΔMAE=0.030 SUVR). All results are listed in Supplementary Tab. 5.

### MRI features contain spatial information predictive of asymmetric tau load

The second tau-PET predicted we predicted was asymmetry in tau load between the two hemispheres, i.e., a Braak I-IV laterality index (LI), specifically in tau-positive participants (assessed with tau-PET). For this task, the *MRI Variables* consistently produced the lowest MSE both alone and in combination with the other feature blocks (Fig. 4a). Support vector regressor (SVR) models resulted in best performance, followed by tree-based (RF and boosting) and elastic net regressors. Details on all 56 combinations and input feature blocks in the ML pipeline can be seen in Supplementary information.

**Fig. 4:**
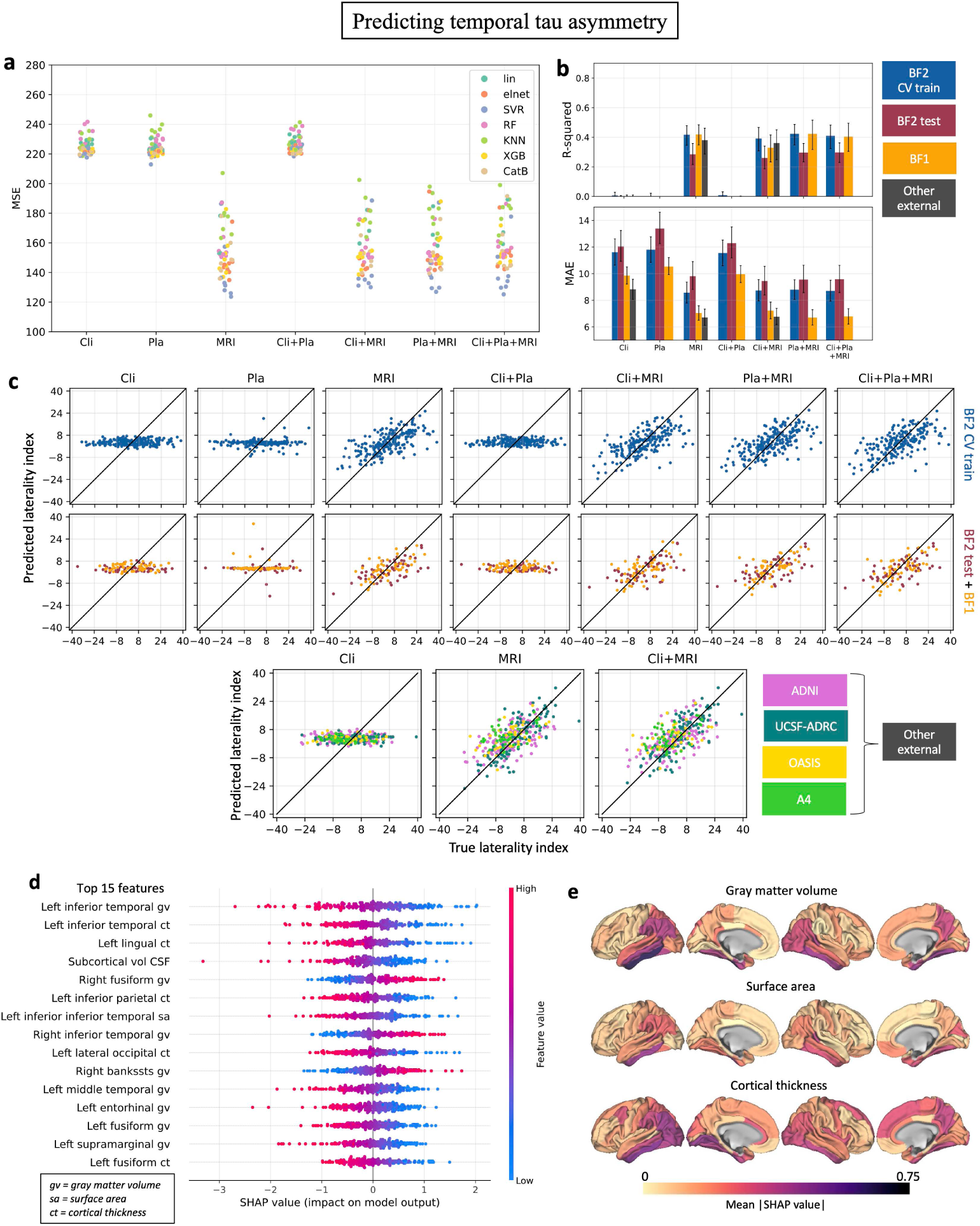
MRI features were most predictive of temporal hemispheric tau asymmetry. **a)** Mean squared error (MSE) of all 56 feature selection + estimator combinations (Fig. 2) for the different input feature blocks (clinical, plasma, MRI, and all combinations) when predicting tau-PET asymmetry in the cross-validated BF2 training set. Each scatter point is colored by estimator. The blocks that included MRI features consistently performed best, always in an SVR model. **b)** R-squared and mean absolute error (MAE) of best pipeline combinations in a) for the cross-validated BF2 training set, BF2 test set, and external test cohorts BF1, ADNI, UCSF-ADRC, OASIS3 and A4. **c)** Scatter plots of true vs predicted laterality index for the best pipeline combinations in a) for the cross-validated BF2 training set and test set, and the other external cohorts. **d)** top SHAP feature contributions in the model including all features, evaluated on the cross-validated BF2 training set. **e)** visualization of the SHAP feature contribution analysis for the MRI FreeSurfer variables. *Abbreviations: Cli (Clinical), Pla (Plasma), MAE (mean absolute error), lin (linear regressor), elnet (elastic net regressor), SVR (support vector regressor), RF (random forest regressor), KNN (K-nearest neighbor regressor), XGB (XGBoost regressor), CatB (CatBoost regressor)*.

The best model for each feature input block (lowest MSE in Fig. 4a, details in Supplementary Tab. 6) was evaluated in the BF2 test set and external cohorts BF1, ADNI, UCSF-ADRC, OASIS3 and A4. Combinations with plasma were only evaluated in BF2 and BF1. R-squared and MAE values varied between cohorts (Fig. 4b and Supplementary Tab. 7), but combinations including the MRI variable block consistently resulted in best performance (R-squared = 0.28-0.42 and MAE = 6.7-9.6 LI). The prediction results are presented in scatter plots, revealing high/consistent generalizability (Fig. 4c).

The top five models within each input block combinations were compared to further explore generalization properties (Supplementary Tab. 6 and Supplementary Fig. 4). The best five models had small variation within input feature blocks for both the BF2 test set and BF1 cohort. No obvious differences in model performance were seen across the five best models within an input feature block. Models including MRI features invariably outperformed models without these features.

SHAP analysis revealed that the most predictive features of tau-PET temporal lobe laterality were temporal lobe MRI gray matter volumes and cortical thicknesses (Fig. 4d and 4e). Reasonably, the model inferred less gray matter in the left hemisphere as predictive of left-laterality, and vice versa (Fig. 4d). For further model transparency, the SHAP interaction plots for the top six features in Fig. 4d are provided in Supplementary Fig. 5.

### Proof-of-concept for a two-step model for clinical use

The translation of predictive models into clinically useful tools is an essential but often overlooked aspect of ML studies in AD. We combined the tasks from the previous two section to create a two-step approach that can allow a clinician to simultaneously evaluate a patient’s tau-PET load and laterality. First, plasma biomarkers would be used to assess if a patient is tau-PET positive. Second, in those classified as tau-PET positive by the blood-biomarkers, an MRI-based algorithm is applied to assess whether the tau pattern is likely to be symmetric or whether tau load is higher in the left or right hemisphere. We reformulated the tasks into classification instead of regression since categorical outcomes often are clearer and easier to interpret, which are important factors in a clinical context.

We simulated such a clinical scenario with a two-step classification task of predicting: 1) the two-label classification task of tau-PET positivity/negativity using plasma variables, and if tau-positive: 2) the three-label classification task of a left-asymmetric/right-asymmetric/symmetric tau phenotype (cutoffs LI=-7.6 and LI=7.3, based on two standard deviations above/below the mean value in the T-negative group) using MRI variables. For this, the ML pipeline was re-run to search for the best performing ML classifiers (note that in the previous sections the estimators were optimized for regression and not classification tasks), which thereafter were evaluated on unseen data using the BF2 test set and BF1. The proposed workflow and results are visualized in Fig. 5, revealing high accuracy between 0.86-0.92 for step one (predicting tau positivity, two label task) using an XGBoost model and substantially better-than-chance performance (accuracy 0.61-0.65) for task two (predicting tau laterality, three label task) using an XGBoost model. This can be further compared against a model always predicting the majority class, which would have yielded an accuracy of 0.65-0.78 for step one, and 0.42-0.45 for step two, based on the class distributions in the test cohorts.

**Fig. 5:**
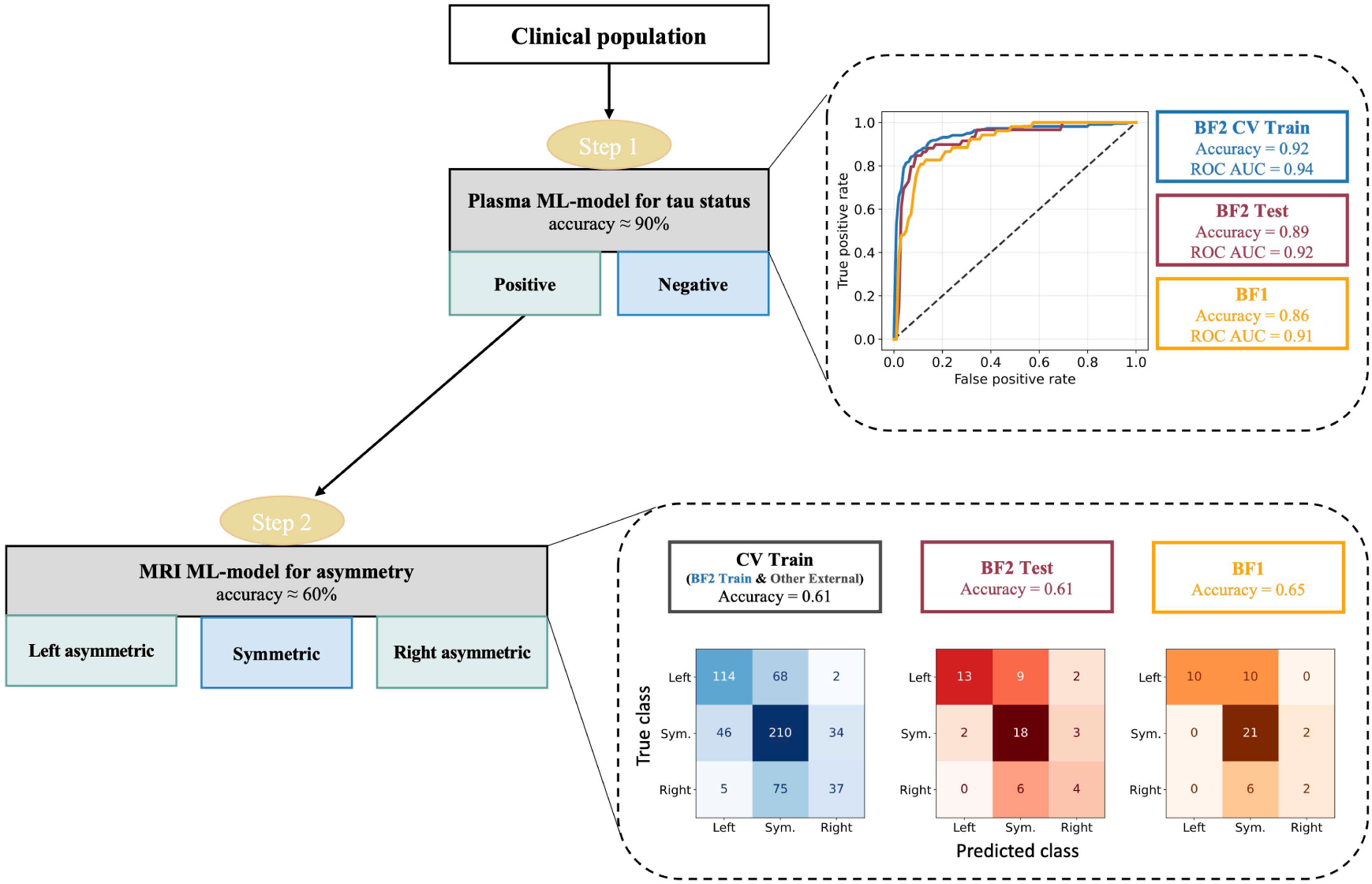
A two-step classification approach could inform about a patient’s predicted temporal tau load and asymmetry from clinically accessible features in ML models. A simulated two-step classification scenario predicting 1) tau-positivity/negativity (load) using plasma variables and if tau-positive: 2) left-asymmetric/symmetric/right-asymmetric using MRI variables. Step one was trained on the full BF2 training set, and step two on tau-PET positive participants from BF2 training set, ADNI, OASIS3, A4 and UCSF-ADRC. Results were evaluated on the cross-validated training set (blue), BF2 test set (red) and external test cohort BF1 (orange). Note that for step two, the training set contained true tau-PET positive participants, while the test sets included predicted positive participants from step one.

Two other potential workflows were also tested: using “clinical + plasma + MRI” variables as input for both steps and using “only MRI” for both steps. The workflow including all feature input blocks performed similar to using plasma variables for task 1 and MRI variables for task 2 (task 1 accuracy: 0.84-0.93, task 2 accuracy: 0.53-0.63), while the “MRI only” workflow performed slightly inferior (task 1 accuracy: 0.82-0.89, task 2 accuracy: 0.46-0.58). See Supplementary Tab. 8 and 9 for details on all workflows.

## Discussion

We address the issue of limited access to tau-PET, a valuable diagnostic tool for Alzheimer’s disease, by exploring the utility of machine learning models and readily available data (clinical, plasma, and structural MRI) to predict important information from tau-PET scans. We formulated two regression tasks: predicting tau-PET SUVR in the temporal lobe and predicting hemispheric asymmetry of temporal lobe tau load in tau-positive participants. We implemented a rigorous ML pipeline that searched for high-performing combinations of feature selection steps and estimators in a combined grid-search and Bayesian optimization set-up. Plasma features (particularly plasma P-tau217) resulted in the best prediction of temporal lobe tau SUVR, while MRI features resulted in best prediction of temporal lobe tau asymmetry. The models were evaluated on unseen test data and independent cohorts, revealing a high degree of model generalizability. We also simulated a two-step model for clinical use, which showed promising results in classifying tau-PET positivity and spatial asymmetry from plasma and MRI variables. This study reveals current potential in predicting tau-PET from more accessible variables, which could dramatically improve diagnosis and prognosis of AD by using scalable, cost-effective methods.

Overcoming the high cost of tau-PET is an important challenge in AD research. Tau-PET has been shown to add clinical information beyond fluid biomarkers, increasing the diagnostic confidence of treating clinicians.^24^ Tau-PET is also used as a screening tool and to monitor treatment effects in clinical trials.^9,11^ Furthermore, results from the TRAILBLAZER-ALZ 2 Randomized Clinical Trial suggested that the amyloid-lowering therapy donanemab provided a greater benefit when initiated at an earlier disease stage (as was indicated by less tau accumulation using tau-PET).^25^ Early therapeutic intervention has also previously been highlighted as key for a more meaningful disease modification in AD.^26,27^ This emphasizes the importance of broad tau-PET accessibility to increase diagnostic confidence, reduce costs in clinical trials and enable effective early treatment in a large population.

Here, we approach this problem with ML models, revealing the potential in combining plasma and MRI features to predict both temporal lobe tau load and spatial patterns. These implications are informative for future studies aimed to improve predictions of tau-PET from more accessible data, for example in studies aimed to synthesize full tau-PET scans using more advanced mathematical models (e.g., deep learning). Such attempts have been made already, but usually high performance has only been reached when using other PET modalities, and thereby not removing the need of a PET scan in the AD diagnostic workflow. Lee et al. were, for example, able to synthesize tau-PET scans with high accuracy using fluorodeoxyglucose (FDG) and amyloid PET, but saw notably inferior performance when using MRI.^28^ Chen et al. showed that full-dose PET images may be predicted from MRI and ultra-low-dose tau-PET images in deep learning models, reducing the radiation but still requiring a PET scan.^29^ Results of our study highlight that combining plasma biomarkers and MRI can be a potential way forward in synthesizing full tau-PET scans without requiring any kind of PET modality as input.

Plasma biomarkers are not yet broadly available, but due to their low cost and high accuracy, they are likely to play an important role in future AD diagnostics. Recent work has even successfully evaluated them in primary care, highlighting their potential for broad clinical use. ^30^ The relationship between plasma biomarkers and tau-PET load has been extensively examined in research studies.^11^ Plasma P-tau217 often shows superior association with temporal lobe tau load compared to other plasma biomarkers (including other P-tau species),^12,31–34^ and even reaches equal or superior performance compared to CSF biomarkers.^35^ This is in line with our results, highlighting the superiority of plasma P-tau217 in predicting temporal lobe tau load at different stages of the disease. Not only did plasma P-tau217 stand out as the main contributing feature, but a linear regression model with only plasma P-tau217 usually outperformed more complex ML models trained without plasma features. Plasma P-tau217 has also been shown efficient when differentiating AD dementia from other neurodegenerative diseases,^32,34,36^ in line with the results presented in Fig 3e. This stratified evaluation suggested that there was little to no gain from adding clinical and MRI variables to a model with only plasma variables in a setting where other neurodegenerative diseases are present (e.g., in primary care or at a memory clinic), as these added variables did not differentiate well between pathological changes due to AD dementia or other dementias. Within the AD dementia group however, a small improvement was seen when adding the clinical and MRI variables, indicating that these features potentially could add more predictive information in a setting where cognitive impairment is very likely due to AD (e.g., a clinical trial). Note that the included clinical variables were aimed to represent highly accessible features, and therefore relatively broadly used and readily administered cognitive tests were selected. More comprehensive or sophisticated cognitive evaluations may improve the differentiation between dementias. Furthermore, plasma P-tau217 has been suggested to relate both to tau and amyloid load in AD.^37^ Upcoming, more tau pathology-specific fluid-based biomarkers (e.g. MTBR-tau243^38^) can likely improve tau-PET prediction even further.

MRI models consistently underestimated the amount of tau in the brain. This may be related to the fact that neurodegeneration measurable with MRI likely occurs after tau has already infiltrated a brain region.^39^ It may also be related to differences in resilience to tau load. Some individuals may be able to harbor tau for long without experiencing atrophy.^40–42^ Moreover, soluble tau phosphorylation patterns detected by plasma provided no predictive information of lateralization in tau load. This may provide some limited evidence against the idea of the hemispheric lateralization being related to differences in the biochemistry of tau. Potential other mechanisms, like for example differences in brain connectivity or genomic factors, could instead be possible underlaying causes of varying lateralization patterns^43^, but this should be further explored in future works.

We proposed a potential two-step clinical platform, which could be of large utility in a clinical population where tau-PET currently is not available. Tau-PET positivity is a well-established marker of the pathological process of AD,^2^ and analyzing spatial tau accumulation patterns enables a more comprehensive understanding of the disease.^18^ Temporal lobe tau laterality has, for example, been suggested to influence behavior and language patterns in AD,^23^ and be linked to faster AD progression,^18^ indicating that spatial information may improve AD disease prognoses in an individualized manner. Here, tau positivity was firstly classified and, if positive, a laterality class of symmetric, left asymmetric or right asymmetric was also predicted. The first classifier was able to discriminate tau-PET positive from negative participants with high accuracy (approximately 90%). The second classifier showed potential with an accuracy of approximately 60%, which can be regarded relatively high considering that this was a multilabel task (always predicting the majority class would result in approximately 40% accuracy). This two-step approach can likely reach even higher accuracies in future studies including upcoming high-performing plasma biomarkers and larger training sets.

The evaluation metrics in this work are well established within the field of ML and are clearly representative of model performance within a cohort. However, when the outcome variable is imbalanced or unequally represented at different parts of the distribution, evaluation metrics can be highly sensitive to cohort properties, potentially influencing the interpretation of model generalizability. When predicting tau-PET asymmetry, lower MAE was, for example, obtained in the external cohorts compared to the cross-validated BF2 (Fig. 4). Usually, ML models are expected to perform similar or worse on external data, as the model unlikely has learned characteristics that are not present in the training set. However, since the external cohorts in this work had a narrower laterality index distribution with fewer extreme left/right asymmetric cases (as seen in Fig. 1), the prediction task becomes easier, which may explain the better performance according to the MAE metric. Furthermore, when predicting tau-PET load (Fig. 3c), the majority of individuals in all cohorts had relatively low tau load and therefore covered a small range of the outcome variable distribution. The expected absolute value of the error hence increased for higher true tau load cases, often making it more informative to compare model performance between cohorts by looking at the scatter plots (Fig. 3c) and/or stratify by diagnostic groups (as was done in Fig. 3e). These results revealed that to improve the models further, additional refinement when predicting load in individuals within the tau-PET positive range is of main importance. This could likely be achieved through larger sample sizes, upcoming more tau-specific fluid-based biomarkers (e.g. MTBR-tau243) and/or more advanced modeling methods.

Limitations of this study include the varying overlap of data between cohorts, e.g., hindering us to evaluate the performance of plasma biomarkers in all external cohorts. Furthermore, the predictive performance of ML estimators is always highly dependent on the data distribution and structures within the available training data. As often is the case in AD cohorts, demographic biases in the data existed, e.g., mainly highly educated, white participants. Future work is needed to assess generalizability to a more diverse population. Nevertheless, this work explores a promising approach to perform tau-PET prediction by incorporating multiple levels of data in machine learning models. Our study highlights both promising and limiting aspects of several clinically accessible variables in this context, with broad relevance for clinical settings and future research.

## Methods

### Study Design

Participants that had undergone thorough expert clinical assessments, tau-PET, MRI and plasma collection were included from two study cohorts: the Swedish BioFINDER-2 (BF2) cohort (enrollment between 2017 and 2022, n=1195, NCT03174938) and the Swedish BioFINDER-1 (BF1) cohort (enrollment between 2010 and 2015, n=147, NCT01208675). All participants in BF2 and BF1 were recruited at Skåne University Hospital or the Hospital of Ängelholm, Sweden. AD dementia was diagnosed according to the Diagnostic and Statistical Manual of Mental Disorders (fifth edition) AD criteria and Aβ-positivity. Further details about BF2 and BF1 have been described in previous works.^34,44^ Detailed demographics are presented in Supplementary Tab. 1.

Participants that had undergone clinical assessments, tau-PET, MRI and were tau-PET positive were included from four additional study cohorts: the Alzheimer’s disease neuroimaging initiative cohort (ADNI; http://adni.loni.usc.edu; n=136), University of California San Francisco Alzheimer’s Disease Research Center cohort (UCSF-ADRC, n=144)^45,46^, the Anti-Amyloid Treatment in Asymptomatic AD cohort (A4, n=45)^47,48^ and Open Access Series of Imaging Studies phase 3 (OASIS-3, n=46)^49^. ADNI is a publicly available cohort, aimed for use in research studies investigating progression to AD. Participants were recruited across centers in North America. The UCSF-ADRC maintains a research clinical cohort, including participants with different neurodegenerative diseases and at various stages of cognitive decline. A4 is a secondary prevention trial in preclinical Alzheimer’s disease, including cognitively unimpaired Aβ-positive participants (based on Aβ-PET). The data used in this work was collected before randomization to treatment arms. OASIS3 is a publicly available cohort study generated by the Knight ADRC and its affiliated studies, including participants at various stages of cognitive decline. Demographics on all cohorts are presented in Supplementary Tab. 1.

### Ethics

All protocols were approved by each cohort’s respective institutional ethical review board, and all participants gave written informed consent.

### Plasma collection and analysis

BF2 and BF1 plasma samples were collected at baseline, handled according to established preanalytical protocols, previously described in detail for both cohorts.^50,51^ All analyses were performed by technicians blinded to all clinical and imaging data. Plasma biomarkers were measured with Eli Lilly assays on a Meso Scale Discovery platform (P-tau217 and P-tau181)^34^ or with Simoa immunoassays developed at the University of Gothenburg (P-tau231, NTA, GFAP and NfL)^52–54^.

### Tau-PET and MR imaging

In BF2, tau-PET was performed using [^18^F]RO948. Standardized uptake value ratio (SUVR) images were created for the 70-90 min post-injection interval using the inferior cerebellar cortex as reference region. In BF1, ADNI, UCSF-ADRC, OASIS3 and A4, tau-PET was performed using [^18^F]flortaucipir. SUVR images were created for the 80–100 post-injection interval using the inferior cerebellar cortex as reference region. T1-weighted volumetric MRI scans were segmented using FreeSurfer v.6.0 (https://surfer.nmr.mgh.harvard.edu/), resulting in native space parcellations of each participant’s brain using the Desikan–Killiany (FreeSurfer) atlas. All FreeSurfer regions (including volumes, surfaces and cortical thicknesses, and total white matter hyperintensities and intracranial volume) were included as input features, allowing for the ML models to find the most informative ones through dimensionality reduction/within the estimator in a data-driven manner. The parcellations were also used to extract mean SUVR values in regions of interest (ROIs) for each participant in native space. All MRI and tau-PET images were processed locally at Lund University, following the same pipeline described previously across studies.^55^

### Tau-PET prediction outcomes

#### Braak I-IV load

A continuous composite variable was created corresponding to Braak I-IV regions^16^, consisting of entorhinal, amygdala, parahippocampal, fusiform and inferior and middle temporal ROIs. The composite was dichotomized according to previously established cutoffs: positivity >1.36 for [^18^F]RO948 and >1.34 for [^18^F]flortaucipir.^5,56^ See Fig. 1c for examples of tau-PET scans with low and high load in this ROI.

#### Laterality Index

Tau-PET asymmetry in the temporal meta-ROI was computed as a laterality index^57,58^,

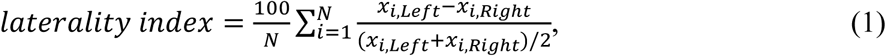

where *x_i_* is the SUVR of brain region *i* and *N* is the total number of brain regions. A negative laterality index represents more tau in the right hemisphere than the left and vice versa. As the index was normalized to the total amount of tau load in the Braak I-IV composite, a more clinically relevant and dispersed distribution was achieved when only including individuals categorized as tau-positive (Fig 1a and 1b). See Fig. 1c for examples of symmetric and asymmetric tau-PET scans in this ROI. For classification tasks, a lower and upper threshold of the composite was computed in the BF2 training cohort as the mean value plus/minus two standard deviations in the tau-negative group: −7.61 and 7.27. Tau-positive participants were divided into three corresponding classes: right-asymmetric (LI < −7.61), left-asymmetric (LI > 7.27) or symmetric (−7.61 < LI < 7.27).

### Machine learning pipeline

A flexible ML pipeline was implemented, designed to handle both different feature types and number of input features for classification and regression tasks. The pipeline is visualized in Fig. 2 and included the following steps:

1. **Standardization.** Performed to get comparable feature scales and numeric adaptation to the estimator’s optimization methods.
2. **Feature selection**. Selecting one of eight different feature selection methods with aim of in a data-driven manner finding the features including relevant predictive information and removing redundant ones, improving chances of generalizability. The eight methods included variations on percentile-based feature selection, recursive feature elimination, and random forest selection, and are described in Supplementary Tab. 2.
3. **Estimator.** Selecting one of seven different estimators with aim of finding the one(s) most suitable for the specific task. The seven methods are described in Supplementary Tab. 2, and include linear models, boosted tree-based methods and support vector machines.
4. **Hyperparameter tuning.** Find appropriate hyperparameters through Bayesian optimization. Bayesian optimization is a technique that efficiently explores the hyperparameter space based on past evaluations. It combines probabilistic models, such as Gaussian processes, with an acquisition function to balance exploration and exploitation, ultimately guiding the search towards promising regions of the hyperparameter space.^59,60^
5. **Evaluation.** Evaluate results on a selected metric relevant to the task at hand. In this work, mean squared error (MSE) was used for all regression tasks and accuracy for all classification tasks.

The full pipeline was trained in a 10-fold cross-validation setting, where all steps were fitted to the training folds and evaluated only for the validation fold. Steps 2) and 3) were conducted in a grid-search setting to compare all 56 combinations of feature selection methods and estimators. The number of samples during Bayesian optimization ranged from 1-50, selected as appropriate depending on the number and range of estimator hyperparameters.

### SHAP analysis

SHapley Additive exPlanations (SHAP) is a method developed by Lundberg et al. for model output explanation, originating from game theory.^61^ It utilizes the Shapley values, which is a measure of how much each feature *x* contributes to the final prediction *f*(*x*), taking interaction effects between features into account (since certain permutations can make features contribute more than the sum of their parts). A local explanatory linear model

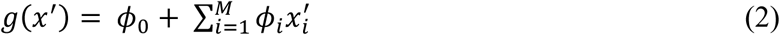

is created that converges to the original model *f*(*x*) when *x*′ ≈ *x*, where *x*′ are simplified input features (binary inclusion/exclusion of a feature), *ϕ*_0_ is the model output with all input features missing, and Shapley values *ϕ* are the coefficients.

### Statistical analysis

When searching for hyperparameter-tuned high-performing ML models, evaluations were performed using 10-fold-cross-validation within the training dataset. When evaluating performance in the BF2 test datasets, the models were first refit on the full training dataset. When evaluating performance in the external test datasets, the models were refit on the full BF2 dataset. All regression models were trained using the BF2 training set. During the two-step classification, step one was trained using the BF2 training set, and step two was trained using tau-PET positive individuals from the BF2 training set, ADNI, OASIS3, A4 and UCSF-ADRC. BF2 test and BF1 were used to test both steps, with only the predicted tau-PET positive individuals from step one evaluated in step two for an as realistic simulation as possible. Regression models were optimized on mean squared error (MSE) and classification models on accuracy. Details on feature selection methods, estimators and metrics can be seen in Supplementary Tab. 2. One-tailed significance testing was performed as bootstrapped difference for a given metric between models. P-values were adjusted for multiple comparisons by the Benjamini–Hochberg false discovery rate method.

### Imputation and missing data

For fair comparison of performance between the different input feature blocks, only participants with complete data for all clinical, plasma and MRI variables were included from BF2. However, no other cohort included an identical set of variables as BF2, so some variables were imputed using a KNN imputer (trained with the training set, number of neighbors = 5) to make the external evaluation possible. Note that if a missing variable was a main contributor during the prediction (assessed in the SHAP analysis), the cohort was not considered sufficient for testing (e.g. missing plasma P-tau217 and/or P-tau181 when predicting temporal lobe tau load, why ADNI, UCSF-ADRC, A4 and OASIS3 were excluded). See Supplementary Tab. 10 for details on variable imputations for each cohort.

## Supporting information

Supplementary machine learning pipeline results for regression task 1 (temporal lobe tau load)

Supplementary machine learning pipeline results for regression task 2 (temporal lobe tau asymmetry)

Supplementary input feature correlation matrix

## Acknowledgements

We would like to express our gratitude to the research volunteers who participated in the studies from which these data were obtained, as well as to their supportive families.

## Author contributions

Conceptualization: LK, JV, IA, KÅ, NMC, OH.

Data curation: LK, OS, RS, APB.

Formal Analysis: LK.

Funding acquisition: JV, NMC, OH.

Project administration: LK.

Resources: OS, ES, NJA, SP, RS, SJ, RLJ, GDR, OH.

Software: LK.

Supervision: JV, IA, KÅ, NMC, OH.

Visualization: LK.

Writing – original draft: LK.

Writing – review and editing: LK, JV, IA, KÅ, OS, JS, RAIB, ES, RO, NJA, HZ, KB, SP, RS, SJ, RLJ, GDR, APB, NMC, OH.

## Data availability

Six different cohorts were used in this work: BF2, BF1, ADNI, UCSF-ADRC, A4 and OASIS3. ADNI, A4 and OASIS3 are publicly available datasets and can be obtained from http://adni.loni.usc.edu/, https://ida.loni.usc.edu/ and https://sites.wustl.edu/oasisbrains/ respectively. The other datasets are not publicly available, but pseudonymized data can be shared with qualified academic researchers upon request (for BF1 and BF2: contact the principal investigator O.H., for UCSF-ADRC: submit a request form at https://memory.ucsf.edu/research-trials/professional/open-science). For BF1 and BF2, data transfer must be performed in agreement with EU legislation regarding general data protection regulation and decisions by the Ethical Review Board of Sweden and Region Skåne.

## Code availability

Code for the analyses can be found in the following GIT repository: https://github.com/DeMONLab-BioFINDER/karlsson-predict-taupet. All analyses were implemented using Python version 3.9. Python dependencies include NumPy^62^, pandas^63^, Matplotlib^64^, Scikit-learn^65^. Note that the flexible ML pipeline is not specific to this work but can be applied for any classification/regression task with spreadsheet data.

## Computational resources

The 56 feature selection + estimator combinations in the ML pipeline were parallelized for time efficiency. Computations were performed on the Bianca Cluster, which is a part the National Academic Infrastructure for Supercomputing in Sweden (NAISS). Bianca is dedicated for analyses of sensitive data, providing 4480 cores in the form of 204 dual CPU (Intel Xeon E5-2630 v3) Huawei XH620 V3 nodes with 128GB memory.

