## Supplementary figures for "A machine learning-based prediction of tau load and distribution in Alzheimer’s disease using plasma, MRI and clinical variables"

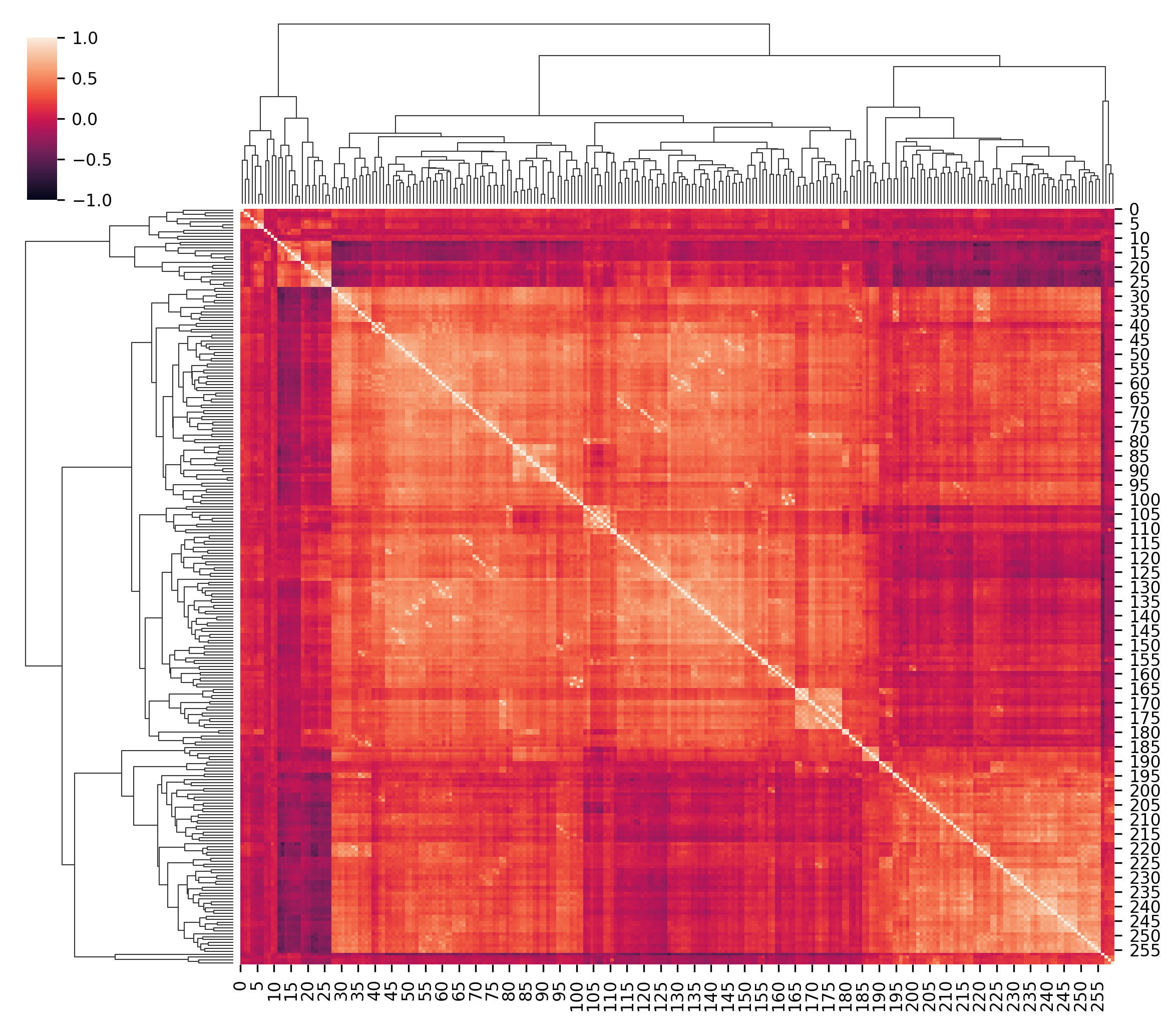

**Supplementary Figure 1: A hierarchically clustered heatmap displaying correlations of all input features.** A correlation matrix between all 260 input features in the BF2 training set, sorted by hierarchical clustering. Certain features covary and may contain very similar information, but the feature space generally provides a diverse range of variability. See supplementary information for mapping of x/y-axis number and feature name, as well as details on exact correlation coefficients.

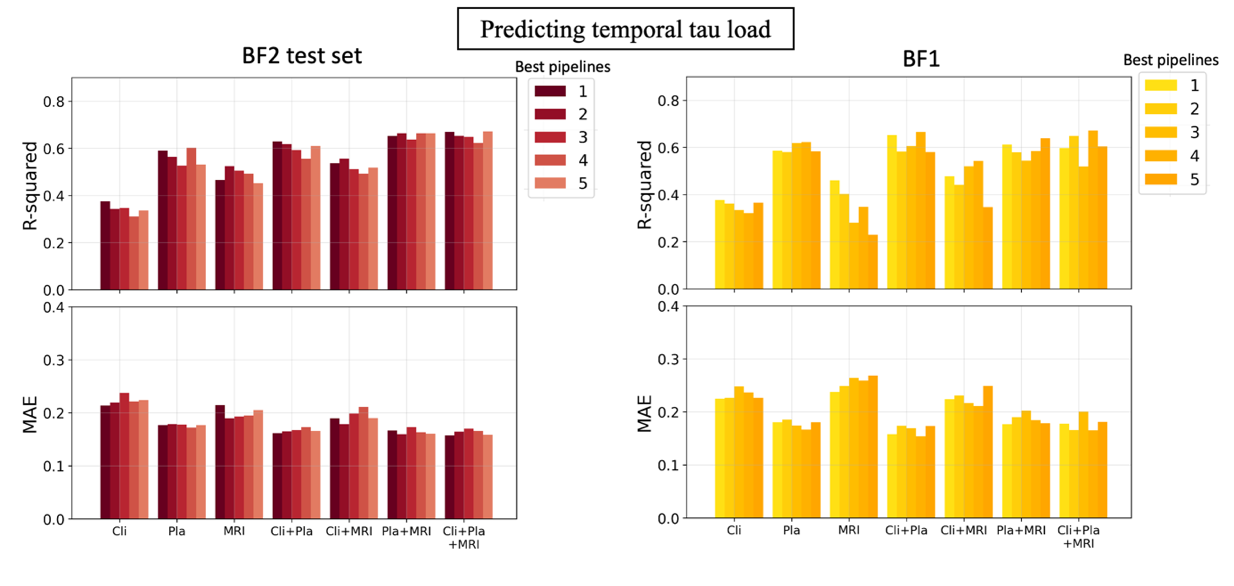

**Supplementary Figure 2: Performance difference between the five top performing models when predicting temporal tau load.** The top five ML models within each input block combinations were compared and evaluated on R-squared and mean absolute error in the BF2 test set and external cohort BF1. All models performed similarly within an input feature block, suggesting there was no clear benefit in selecting either one of the top performing models. Differences between feature input blocks remained.

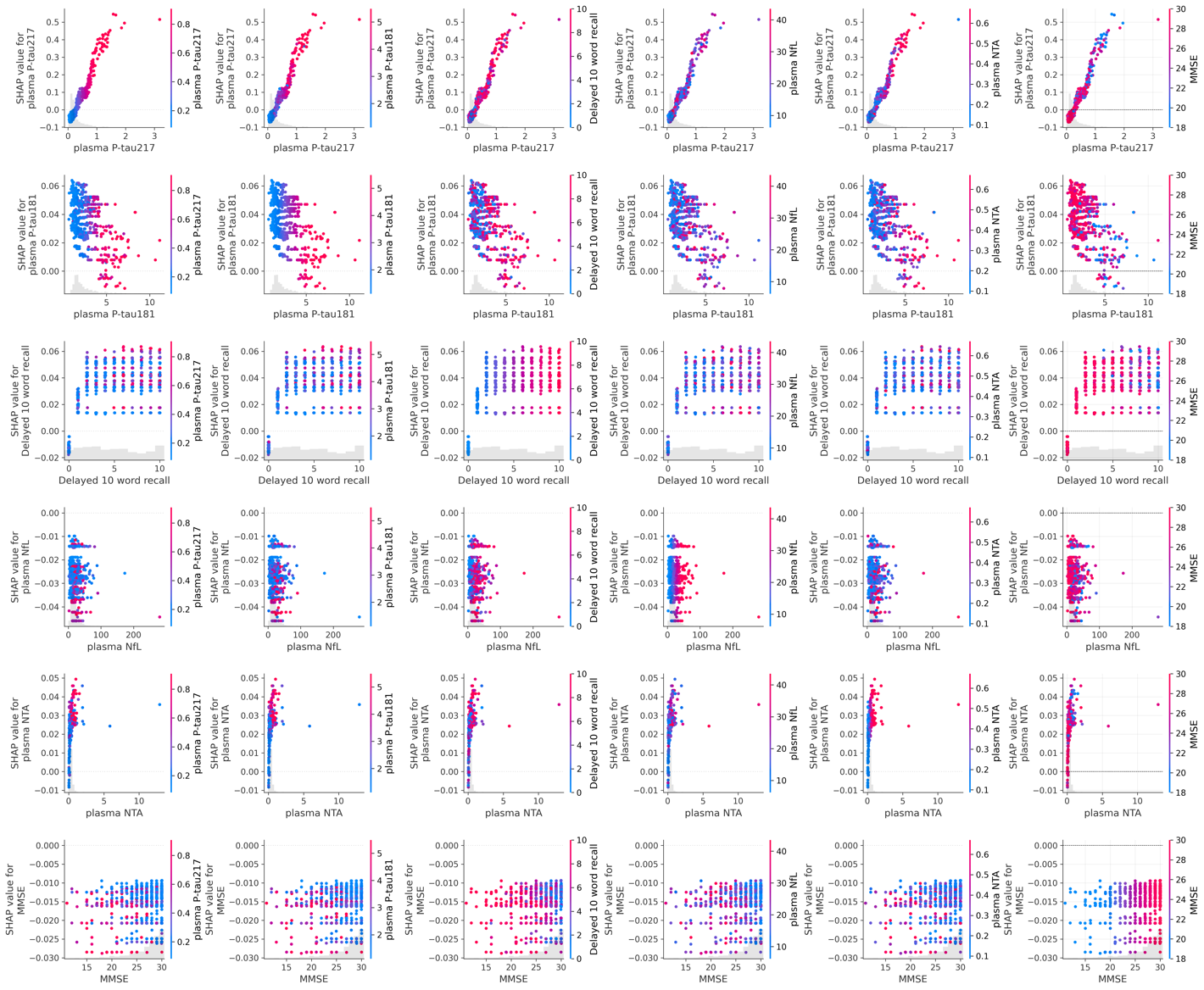

**Supplementary Figure 3: SHAP interaction plots for the top six features when predicting temporal tau load.** SHAP interaction analysis between the top six features for the best performing model (same as in Fig. 3d).

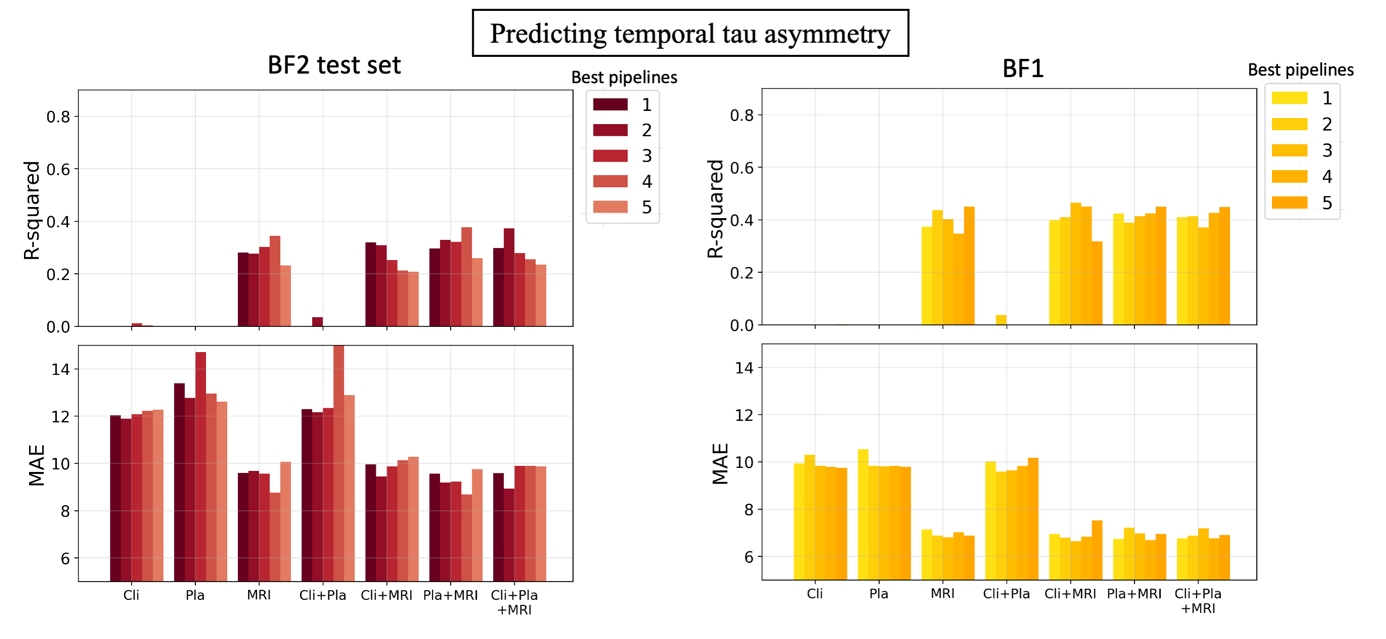

**Supplementary Figure 4: Performance difference between the five top performing models when predicting temporal tau asymmetry.** The top five ML models within each input block combinations were compared and evaluated on R-squared and mean absolute error in the BF2 test set and external cohort BF1. All models performed similarly within an input feature block, suggesting there was no clear benefit in selecting either one of the top performing models. Differences between feature input blocks remained.

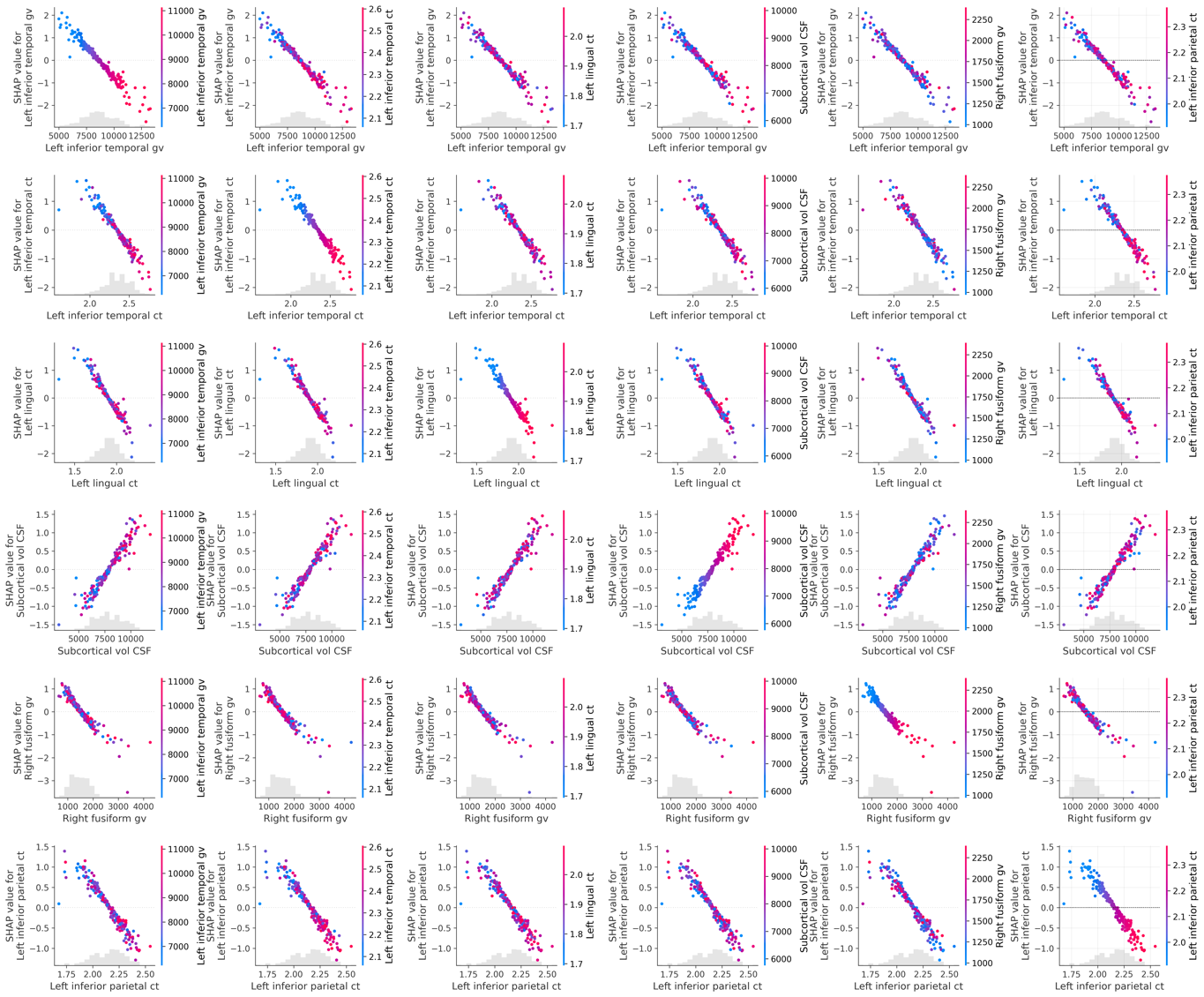

**Supplementary Figure 5: SHAP interaction plots for the top six features when predicting temporal tau asymmetry.** SHAP interaction analysis between the top six features for the best performing model (same as in Fig. 4d).
