## Supplementary tables for "A machine learning-based prediction of tau load and distribution in Alzheimer’s disease using plasma, MRI and clinical variables"

**Supplementary Table 1: Cohort demographics.** Characteristics of all cohorts used in this work. Tau positive participants (Braak I-IV load > 1.36 in BF2 and Braak I-IV load > 1.34 in all other cohorts) were divided into three classes: right asymmetric (LI < -7.61), left asymmetric (LI > 7.27) or symmetric (-7.61 < LI < 7.27). Abbreviations: laterality index (LI), mini mental state examination (MMSE), positron emission tomography (PET), normal cognition (NC), subjective cognitive decline (SCD), mild cognitive impairment (MCI), standardized uptake value ratio (SUVR),

*missing data for some participants

|  | **BioFINDER-2** | | **BioFINDER-1** | **ADNI (only T+)** | **UCSF-ADRC (only T+)** | **OASIS (only T+)** | **A4  (only T+)** |
| --- | --- | --- | --- | --- | --- | --- | --- |
|  | *Train* | *Test* |  |  |  |  |  |
| **n** | 948 | 247 | 147 | 136 | 144 | 46 | 45 |
| **Age [years]*** | 67.5 (12.5) | 67.7 (12.4) | 72.8 (7.37) | 72.8 (6.90) | 63.5 (8.21)  *n=109 | 74.7 (6.75) | 72.7 (4.75) |
| **Sex male (%)*** | 486 (51%) | 132 (53%) | 82 (56%) | 64 (47%) | 44 (40%)  *n=109 | 23 (50%) | 16 (35%) |
| **Education [years]*** | 12.7 (3.79) | 12.7 (3.75) | 12.2 (3.60) | 15.9 (2.37) | 17.0 (2.83)  *n=90 | 16.0 (2.61) | 16.7 (2.66) |
| **MMSE*** | 26.8 (3.77) | 26.8 (3.60) | 25.9 (4.44) | 25.3 (4.09) | 21.3 (5.54)  *n=92 | 25.8 (4.01) | 28.2 (1.41) |
| **ADAS delayed word recall** | 4.82 (3.21) | 4.70 (3.16) | 4.97 (3.38) | 6.24 (3.18) | - | - | - |
| ***APOE* ε4 carrier* (%)** | 481 (51%) | 126 (51%) | 84 (57%) | 77/115* (70%) | 44/82* (54%) | 28 (61%) | - |
| **Clinical Diagnosis*** |  |  |  |  |  |  |  |
| *NC* | 321 | 83 | 54 | 24 | - | 17 | 45 |
| *SCD* | 153 | 33 | 9 | - | - | - | - |
| *MCI* | 236 | 77 | 25 | 49 | 16 | - | - |
| *Dementia* | 232 | 53 | 41 | 61 | 61 | 29 | - |
| *Other* | 6 | 1 | 18 | - | 30 | - | - |
| **Tau-PET and plasma biomarkers** | | | | | | | |
| ***Tau negative*** | | | | | | | |
| n | 728 | 188 | 95 | - | - | - | - |
| Braak I-IV load [SUVR] | 1.15 (0.090) | 1.16 (0.100) | 1.16 (0.067) | - | - | - | - |
| Braak I-IV Laterality Index | -0.169 (3.72) | -0.118 (3.57) | 0.734 (2.98) | - | - | - | - |
| Plasma P-tau217 [pg/ml] | 0.198 (0.159) | 0.205 (0.144) | 0.259 (0.135) | - | - | - | - |
| Plasma P-tau181 [pg/ml] | 1.90 (0.805) | 2.01 (0.901) | 2.50 (2.89) | - | - | - | - |
| ***Tau positive*** | | | | | | | |
| n | 220 | 59 | 52 | 136 | 144 | 46 | 45 |
| Braak I-IV load [SUVR] | 2.04 (0.575) | 1.96 (0.545) | 1.87 (0.400) | 1.67 (0.335) | 2.05 (0.436) | 1.67 (0.300) | 1.45 (0.115) |
| Braak I-IV Laterality Index | 2.24 (14.9) | 3.05 (16.5) | 2.86 (11.6) | 1.10 (11.1) | 3.63 (11.0) | 1.31 (12.4) | -1.67 (8.52) |
| Plasma P-tau217 [pg/ml] | 0.687 (0.332) | 0.688 (0.422) | 0.682 (0.381) | - | - | - | - |
| Plasma P-tau181 [pg/ml] | 4.13 (1.65) | 4.10 (2.18) | 4.69 (2.47) | - | - | - | - |
| Symmetric | 93 | 21 | 22 | 69 | 75 | 9 | 27 |
| Left asymmetric | 80 | 26 | 20 | 37 | 49 | 11 | 7 |
| Right asymmetric | 47 | 12 | 10 | 30 | 20 | 26 | 11 |

**Supplementary Table 2: Description of the feature selection methods, machine learning estimators and metrics used to evaluate classification and regression tasks.**

Abbreviations: True positives (TP), True negatives (TN), False positives (FP), False negatives (FN).

| **Feature selection method** | **Description** |
| --- | --- |
| None | No feature selection – all input features used. |
| 10% F-statistic | Select the top 10% features with highest i) ANOVA F-value (classification) or ii) univariate linear regression F-value (regression) computed between feature and outcome variable. |
| 50% F-statistic | Select the top 50% features with highest i) ANOVA F-value (classification) or ii) univariate linear regression F-value (regression) computed between feature and outcome variable. |
| 10% mutual info | Select the top 10% features with highest Mutual information (MI) score, a nonparametric method based on entropy estimation.^1^ |
| 50% mutual info | Select the top 50% features with highest Mutual information (MI) score, a nonparametric method based on entropy estimation.^1^ |
| 10% RFE | Recursive feature elimination, recursively pruning the least informative features in a Random Forest classifier/regressor until 10% left. |
| 50% RFE | Recursive feature elimination, recursively pruning the least informative features in a Random Forest classifier/regressor until 50% left. |
| 95% cum RF | Sorts the feature importance values from a Random Forest classifier/regressor and selects those needed to create a cumulative sum representing 95% of the total sum. |
| **Machine learning estimator** |  |
| Linear | Classification: Logistic regression.  Regression: Linear regression. |
| ElasticNet | Classification: Logistic regression with both L1 and L2 terms as regularization.  Regression: Linear regression with both L1 and L2 terms as regularization. |
| SVC/SVR | Support vector classifier: classify data by finding an optimal hyperplane that maximally separates different classes.  Support vector regressor: predict continuous target variables by finding an optimal hyperplane that fits the data. |
| RF | Random Forest: ensemble learning algorithm that combines multiple decision trees to make predictions by aggregating the votes of individual trees. |
| KNN | K-nearest neighbor: makes a prediction by averaging the outcome of the k nearest neighbors in the feature space. |
| XGBoost | XGBoost: a gradient boosting algorithm that sequentially combines weak learners to create an ensemble model with high predictive performance.^2^ |
| CatBoost | CatBoost: a gradient boosting algorithm aimed to be superior during handling of categorical features and without the need for explicit data preprocessing.^3^ |
| **Metrics classifier** |  |
| Accuracy | (TP+TN)/(TP+FP+TN+FN) |
| Precision | TP/(TP+FP) |
| Recall | TP/(TP+FN) |
| AUC | Area under the Receiver Operator Characteristic (ROC) curve, a probability curve that displays the relationship between the false positive rate and true positive rate. |
| **Metrics regressor** |  |
| MSE | Mean squared error: average squared difference between the estimated value and the true value. |
| MAE | Mean absolute error: absolute difference between the estimated value and the true value. |
| R-squared | The proportion of variance in the true value that can be explained by the predicted variable. |

**Supplementary Table 3: The best models for the two tasks of predicting temporal tau load and asymmetry for all seven input feature blocks.** Specifics of mean squared error (MSE) of the best feature selection method, estimator, and estimator parameters after running the flexible machine learning pipeline for the seven input feature blocks (feature combinations). The table includes the two regression tasks of predicting temporal tau lad and asymmetry, and these models were later used to evaluate performance in the test sets.

| Predict tau temporal load | | | | |
| --- | --- | --- | --- | --- |
| **Feature combination** | **MSE** | **Feature selection** | **Estimator** | **Estimator params** |
| Clinical | 0.1443 | None | CatBoost | depth = 3 learning rate = 0.02149 number of estimators = 291 |
| Plasma | 0.07235 | 95% cum RF | CatBoost | depth = 10 learning rate = 0.01050 number of estimators = 471 |
| MRI | 0.1064 | 50% F-statistic | SVR | kernel = rbf C = 1.585 |
| Clinical + Plasma | 0.06876 | None | CatBoost | depth = 5 learning rate = 0.02955 number of estimators = 299 |
| Clinical + MRI | 0.09641 | None | CatBoost | depth = 4 learning rate = 0.04120 number of estimators = 500 |
| Plasma + MRI | 0.06344 | 10% RFE | CatBoost | depth = 7 learning rate = 0.04290 number of estimators = 443 |
| Clinical + Plasma + MRI | 0.06088 | 95% cum RF | CatBoost | depth = 5 learning rate = 0.04194 number of estimators = 500 |
| Predict tau temporal asymmetry | | | | |
| Clinical | 217.6 | None | SVR | kernel = rbf C = 0.9495 |
| Plasma | 212.9 | 50% mutual info | SVR | kernel = poly C = 0.5510 |
| MRI | 123.7 | 50% RFE | SVR | kernel = rbf C = 29.43 |
| Clinical + Plasma | 218.4 | None | SVR | kernel = rbf C = 1.177 |
| Clinical + MRI | 130.1 | 50% RFE | SVR | kernel = rbf C = 1000 |
| Plasma + MRI | 127.0 | 50% F-statistic | SVR | kernel = rbf C = 30.51 |
| Clinical + Plasma + MRI | 125.3 | 50% F-statistic | SVR | kernel = rbf C = 1000 |

**Supplementary Table 4: The top five models when predicting temporal tau load for all seven input feature blocks, evaluated in the cross-validated BF2 training set, BF2 test set and external test cohort BF1.** R-squared and mean absolute error (MAE) of the top five feature selection methods, estimators, and estimator parameters after running the flexible machine learning pipeline for the seven input feature blocks (feature combinations).

| **Feature combination** | **Feature selection** | **Estimator** | **R-squared CV BF2 train** | **MAE CV BF2 train** | **R-squared BF2 test** | **MAE BF2 test** | **R-squared BF1** | **MAE BF1** |
| --- | --- | --- | --- | --- | --- | --- | --- | --- |
| Clinical | **None** | **CatBoost** | **0.3605** | **0.2247** | **0.3753** | **0.2135** | **0.3777** | **0.2249** |
|  | None | XGBoost | 0.3397 | 0.2304 | 0.3426 | 0.2195 | 0.361 | 0.2266 |
|  | 95% cum RF | ElasticNet | 0.3261 | 0.2489 | 0.3468 | 0.2374 | 0.3338 | 0.2479 |
|  | 50% mutual info | RF | 0.3214 | 0.2330 | 0.3103 | 0.2217 | 0.3207 | 0.2366 |
|  | 50% RFE | CatBoost | 0.3376 | 0.2312 | 0.3361 | 0.2236 | 0.3651 | 0.2265 |
| Plasma | **95% cum RF** | **CatBoost** | **0.6322** | **0.1625** | **0.5898** | **0.1764** | **0.5864** | **0.1804** |
|  | None | CatBoost | 0.6485 | 0.1598 | 0.5638 | 0.1790 | 0.5799 | 0.1850 |
|  | 95% cum RF | SVR | 0.6389 | 0.1603 | 0.5260 | 0.1778 | 0.6186 | 0.1741 |
|  | 95% cum RF | RF | 0.6475 | 0.1616 | 0.6023 | 0.1719 | 0.6228 | 0.1667 |
|  | None | SVR | 0.6524 | 0.1563 | 0.5314 | 0.1768 | 0.5834 | 0.1806 |
| MRI | **50% F-statistic** | **SVR** | **0.5239** | **0.2166** | **0.4660** | **0.2143** | **0.4601** | **0.2379** |
|  | 50% RFE | CatBoost | 0.5033 | 0.2033 | 0.5245 | 0.1894 | 0.4026 | 0.2488 |
|  | 50% F-statistic | CatBoost | 0.4882 | 0.2071 | 0.5050 | 0.1927 | 0.2798 | 0.2639 |
|  | 95% cum RF | CatBoost | 0.5219 | 0.2028 | 0.4930 | 0.1951 | 0.3483 | 0.2589 |
|  | 50% mutual info | CatBoost | 0.5026 | 0.2040 | 0.4515 | 0.2055 | 0.2292 | 0.2684 |
| Clinical + Plasma | **None** | **CatBoost** | **0.6965** | **0.1482** | **0.6297** | **0.1615** | **0.6522** | **0.1579** |
|  | 95% cum RF | CatBoost | 0.6832 | 0.1490 | 0.6181 | 0.1649 | 0.5819 | 0.1737 |
|  | None | XGBoost | 0.6496 | 0.1558 | 0.5934 | 0.1675 | 0.6054 | 0.1691 |
|  | 50% mutual info | CatBoost | 0.6748 | 0.1524 | 0.5553 | 0.1728 | 0.6650 | 0.1541 |
|  | 50% RFE | CatBoost | 0.6801 | 0.1507 | 0.6103 | 0.1657 | 0.5801 | 0.1733 |
| Clinical + MRI | **None** | **CatBoost** | **0.5539** | **0.1910** | **0.5360** | **0.1895** | **0.4781** | **0.2238** |
|  | 50% F-statistic | CatBoost | 0.5353 | 0.1942 | 0.5571 | 0.1784 | 0.4415 | 0.2306 |
|  | 50% F-statistic | SVR | 0.5614 | 0.2030 | 0.5125 | 0.1986 | 0.5207 | 0.2167 |
|  | 50% mutual info | SVR | 0.5400 | 0.2085 | 0.4932 | 0.2113 | 0.5424 | 0.2113 |
|  | 50% F-statistic | XGBoost | 0.5563 | 0.1948 | 0.5177 | 0.1902 | 0.3468 | 0.2489 |
| Plasma + MRI | **10% RFE** | **CatBoost** | **0.6959** | **0.1489** | **0.6497** | **0.1660** | **0.6440** | **0.1692** |
|  | 95% cum RF | CatBoost | 0.7121 | 0.1529 | 0.6640 | 0.1599 | 0.5795 | 0.1898 |
|  | None | CatBoost | 0.6920 | 0.1570 | 0.6373 | 0.1727 | 0.5432 | 0.2028 |
|  | 50% RFE | CatBoost | 0.6975 | 0.1521 | 0.6644 | 0.1629 | 0.5838 | 0.1842 |
|  | 50% mutual info | CatBoost | 0.6987 | 0.1490 | 0.6643 | 0.1608 | 0.6384 | 0.1789 |
| Clinical + Plasma + MRI | **95% cum RF** | **CatBoost** | **0.7170** | **0.1458** | **0.6836** | **0.1577** | **0.6618** | **0.1642** |
|  | 50% RFE | CatBoost | 0.7227 | 0.1473 | 0.6528 | 0.1645 | 0.6495 | 0.1658 |
|  | 50% RFE | XGBoost | 0.7144 | 0.1512 | 0.6486 | 0.1703 | 0.5191 | 0.2002 |
|  | 50% F-statistic | CatBoost | 0.7158 | 0.1468 | 0.6229 | 0.1659 | 0.6707 | 0.1654 |
|  | 10% mutual info | CatBoost | 0.7031 | 0.1476 | 0.6717 | 0.1584 | 0.6043 | 0.1811 |

**Supplementary Table 5: Comparison of the best models for each input feature block combination against a simple linear regression model with plasma P-tau217 only during predicting of temporal tau load.** R-squared and mean absolute error (MAE) of the top performing ML models. P-values represent a comparison against the simple linear regression model using P-tau217 only, which was assessed with bootstrapping and FDR-corrected. Significant differences are highlighted in bold.

| **Feature combination** | **Feature selection** | **Estimator** | **R-squared BF2 test (P-value, FDR corrected)** | **MAE BF2 test (P-value, FDR corrected)** | **R-squared BF1 (P-value, FDR corrected)** | **MAE BF1**  **(P-value, FDR corrected)** |
| --- | --- | --- | --- | --- | --- | --- |
| Plasma p-tau217 only | | **LinReg** | 0.500 | 0.197 | 0.593 | 0.188 |
| Clinical | **None** | **CatBoost** | 0.3753 (0.92) | 0.2135 (0.89) | 0.3777 (0.98) | 0.2249 (0.98) |
| Plasma | **95% cum RF** | **CatBoost** | 0.5898 (0.45) | 0.1764 (0.12) | 0.5864 (0.98) | 0.1804 (0.38) |
| MRI | **50% F-statistic** | **SVR** | 0.4660 (0.74) | 0.2143 (0.89) | 0.4601 (0.98) | 0.2379 (0.98) |
| Clinical + Plasma | **None** | **CatBoost** | **0.6297 (0.037)** | **0.1615 (0.0023)** | 0.6522 (0.42) | **0.1579 (0.007)** |
| Clinical + MRI | **None** | **CatBoost** | 0.5360 (0.45) | 0.1895 (0.44) | 0.4781 (0.98) | 0.2238 (0.98) |
| Plasma + MRI | **10% RFE** | **CatBoost** | **0.6497** (**0.021)** | **0.1660 (0.0023)** | 0.6440 (0.29) | 0.1692 (0.25) |
| Clinical + Plasma + MRI | **95% cum RF** | **CatBoost** | **0.6836 (0.007)** | **0.1577 (0.0023)** | 0.6618 (0.21) | 0.1642 (0.22) |

**Supplementary Table 6: The top five models when predicting temporal tau asymmetry for all seven input feature blocks, evaluated in the cross-validated BF2 training set, BF2 test set and external test cohort BF1.** R-squared and mean absolute error (MAE) of the top five feature selection methods, estimators, and estimator parameters after running the flexible machine learning pipeline for the seven input feature blocks (feature combinations).

| **Feature combination** | **Feature selection** | **Estimator** | **R-squared CV BF2 train** | **MAE CV BF2 train** | **R-squared BF2 test** | **MAE BF2 test** | **R-squared BF1** | **MAE BF1** |
| --- | --- | --- | --- | --- | --- | --- | --- | --- |
| Clinical | **None** | **SVR** | **0.0068** | **11.6521** | **-0.0093** | **12.0388** | **-0.0307** | **9.9309** |
|  | None | KNN | -0.0034 | 11.64 | -0.0372 | 11.8885 | -0.0552 | 10.295 |
|  | 10% mutual info | SVR | -0.0285 | 11.8008 | 0.0107 | 12.0723 | -0.0056 | 9.8274 |
|  | 95% cum RF | CatBoost | -0.0041 | 11.6689 | 0.004 | 12.2224 | -0.0475 | 9.7891 |
|  | 50% mutual info | CatBoost | -0.0133 | 11.7388 | -0.0039 | 12.277 | 0.0016 | 9.7411 |
| Plasma | **50% mutual info** | **SVR** | **-0.0029** | **11.6881** | **-0.1392** | **13.388** | **-0.275** | **10.5314** |
|  | 50% F-statistic | CatBoost | -0.0456 | 11.9007 | -0.0845 | 12.7668 | -0.0282 | 9.8337 |
|  | 95% cum RF | SVR | -0.024 | 11.7943 | -1.2968 | 14.7002 | -0.0052 | 9.8075 |
|  | 95% cum RF | XGBoost | -0.0246 | 11.8172 | -0.0699 | 12.9479 | -0.0644 | 9.8277 |
|  | 10% mutual info | XGBoost | -0.0206 | 11.8505 | -0.0177 | 12.6162 | -0.0317 | 9.7908 |
| MRI | **50% RFE** | **SVR** | **0.4055** | **8.7347** | **0.2798** | **9.6** | **0.3738** | **7.1467** |
|  | 95% cum RF | SVR | 0.4035 | 8.7664 | 0.2766 | 9.67 | 0.4361 | 6.8738 |
|  | 50% F-statistic | SVR | 0.3952 | 8.835 | 0.3026 | 9.5595 | 0.4018 | 6.8008 |
|  | 10% RFE | SVR | 0.3963 | 8.6627 | 0.344 | 8.7586 | 0.3474 | 7.0199 |
|  | None | SVR | 0.4056 | 8.7897 | 0.232 | 10.0648 | 0.449 | 6.8749 |
| Clinical + Plasma | **None** | **SVR** | **0.0014** | **11.5395** | **-0.0293** | **12.2939** | **-0.0366** | **10.012** |
|  | None | CatBoost | -0.0359 | 11.8044 | 0.0345 | 12.1631 | 0.0373 | 9.588 |
|  | 50% mutual info | CatBoost | -0.0067 | 11.6703 | -0.0018 | 12.3421 | -0.0012 | 9.643 |
|  | 95% cum RF | SVR | -0.081 | 11.9345 | -20.3103 | 21.9158 | -0.0096 | 9.8166 |
|  | None | ElasticNet | -0.0198 | 11.6629 | -0.1122 | 12.8772 | -0.0582 | 10.1683 |
| Clinical + MRI | **50% RFE** | **SVR** | **0.4102** | **8.7678** | **0.3195** | **9.9504** | **0.3972** | **6.9511** |
|  | 50% F-statistic | SVR | 0.4138 | 8.7802 | 0.3084 | 9.4322 | 0.4097 | 6.7949 |
|  | 95% cum RF | SVR | 0.4076 | 8.6881 | 0.252 | 9.8673 | 0.4643 | 6.6326 |
|  | None | SVR | 0.367 | 9.0013 | 0.2127 | 10.137 | 0.4504 | 6.8216 |
|  | 50% mutual info | SVR | 0.3334 | 9.4205 | 0.2069 | 10.2865 | 0.3176 | 7.5218 |
| Plasma + MRI | **50% F-statistic** | **SVR** | **0.4294** | **8.6086** | **0.2963** | **9.5598** | **0.4231** | **6.7395** |
|  | 50% RFE | SVR | 0.3864 | 8.9489 | 0.329 | 9.185 | 0.3888 | 7.2106 |
|  | 95% cum RF | SVR | 0.3653 | 8.9795 | 0.3213 | 9.2252 | 0.413 | 6.9714 |
|  | 10% RFE | SVR | 0.4088 | 8.6116 | 0.3779 | 9.2252 | 0.4244 | 6.6909 |
|  | None | SVR | 0.3552 | 9.1729 | 0.2591 | 9.7485 | 0.4491 | 6.9525 |
| Clinical + Plasma + MRI | **50% F-statistic** | **SVR** | **0.436** | **8.5507** | **0.297** | **9.5851** | **0.4102** | **6.7602** |
|  | 10% RFE | SVR | 0.3483 | 9.1206 | 0.372 | 8.9268 | 0.4133 | 6.8741 |
|  | 50% RFE | SVR | 0.3483 | 9.1615 | 0.2778 | 9.8886 | 0.3705 | 7.1884 |
|  | 95% cum RF | SVR | 0.37 | 9.0147 | 0.2548 | 9.8887 | 0.4266 | 6.7582 |
|  | None | SVR | 0.377 | 9.0149 | 0.235 | 9.8643 | 0.4486 | 6.9025 |

| **Feature combination** | **R-squared ADNI** | **MAE ADNI** | **R-squared UCSF-ADRC** | **MAE UCSF-ADRC** | **R-squared OASIS** | **MAE OASIS** | **R-squared A4** | **MAE A4** | **R-squared other external** | **MAE other external** |
| --- | --- | --- | --- | --- | --- | --- | --- | --- | --- | --- |
| Clinical | -0.05858 | 8.7762 | -0.0021 | 9.0044 | -0.04203 | 9.0972 | -0.3701 | 8.4446 | -0.0234 | 8.8173 |
| MRI | 0.3176 | 7.0156 | 0.58036 | 5.6901 | 0.2553 | 7.3994 | -0.1121 | 7.1343 | 0.3800 | 6.7096 |
| Clinical + MRI | 0.2856 | 7.2116 | 0.5366 | 5.8740 | 0.2593 | 7.6289 | -0.1994 | 7.3819 | 0.3608 | 6.7639 |

**Supplementary Table 7: Model performance on external test cohorts ADNI, UCSF-ADRC, OASIS and A4 when predicting temporal tau asymmetry.** R-squared and mean absolute error (MAE) of the top feature selection methods, estimators, and estimator parameters in Table S6. Other external represents the score when combining the prediction in all three cohorts.

**Supplementary Table 8: Predicting tau positivity (task 1) in the two-step classification task.** Details on feature selection and estimator used to evaluate accuracy, precision, recall and AUC for the binary classification task of tau PET positivity. Evaluation was performed on feature combinations plasma, MRI and clinical+plasma+MRI. Best scores are highlighted in bold.

| **Feature combination** | **Feature selection** | **Estimator** | **CV BF2 train** | | | | **BF2 test** | | | | **BF1** | | | |
| --- | --- | --- | --- | --- | --- | --- | --- | --- | --- | --- | --- | --- | --- | --- |
|  |  |  | Accuracy | Precision | Recall | AUC | Accuracy | Precision | Recall | AUC | Accuracy | Precision | Recall | AUC |
| Step 1: Plasma | None | XGBoost | 0.920 | 0.840 | 0.809 | 0.942 | 0.887 | 0.772 | 0.746 | 0.922 | **0.857** | **0.804** | **0.788** | **0.907** |
| Step 1: MRI | 50% F-statistic | SVC | 0.873 | 0.794 | 0.614 | 0.892 | 0.887 | 0.782 | 0.729 | 0.894 | 0.823 | 0.795 | 0.673 | 0.873 |
| Step 1: Clinical + Plasma + MRI | None | CatBoost | **0.927** | **0.854** | **0.827** | **0.958** | **0.899** | **0.793** | **0.780** | **0.925** | 0.844 | 0.784 | 0.769 | 0.891 |

**Supplementary Table 9: Predicting symmetric, left asymmetric or right asymmetric (task 2) in the two-step classification task.** Details on feature selection and estimator used to evaluate accuracy, precision, recall and AUC in the multilabel asymmetry classification task. For the test sets, evaluation was performed on the subsample of individuals classified as tau PET positive in step 1.
* Step 2: Note that for the combination “Clinical + Plasma + MRI”, only the BF2 training set was used (and not other external as well, as these cohorts did not include plasma features).

| **Feature combination** | **Feature selection + Estimator** | **Class** | **CV BF2 train +**  **Other External*** | | | **BF2 test** | | | **BF1** | | |
| --- | --- | --- | --- | --- | --- | --- | --- | --- | --- | --- | --- |
|  |  |  | Accuracy | Precision | Recall | Accuracy | Precision | Recall | Accuracy | Precision | Recall |
| Step 1: Plasma  Step 2:  MRI | None + XGBoost | Left Asym. | 0.611 | 0.691 | 0.620 | 0.614 | 0.867 | 0.542 | 0.647 | 1.00 | 0.500 |
|  |  | Sym. |  | 0.595 | 0.724 |  | 0.545 | 0.783 |  | 0.568 | 0.913 |
|  |  | Right Asym. |  | 0.507 | 0.316 |  | 0.444 | 0.400 |  | 0.500 | 0.250 |
| Step 1: MRI  Step 2: MRI | None + XGBoost | Left Asym. | 0.607 | 0.663 | 0.576 | 0.564 | 0.813 | 0.542 | 0.568 | 0.75 | 0.353 |
|  |  | Sym. |  | 0.584 | 0.734 |  | 0.500 | 0.727 |  | 0.543 | 0.905 |
|  |  | Right Asym. |  | 0.606 | 0.342 |  | 0.286 | 0.222 |  | 0 | 0 |
| Step 1: Clinical + Plasma + MRI  Step 2*: Clinical + Plasma + MRI | 50% RFE + SVC | Left Asym. | 0.571 | 0.644 | 0.662 | 0.534 | 0.846 | 0.440 | 0.627 | 1.0 | 0.263 |
|  |  | Sym. |  | 0.540 | 0.642 |  | 0.463 | 0.864 |  | 0.561 | 0.958 |
|  |  | Right Asym. |  | 0.500 | 0.273 |  | 0.250 | 0.091 |  | 0.800 | 0.500 |

**Supplementary Table 10: Imputed data in each external test cohort.** The features were imputed due to missingness. Imputation was performed with a KNN imputer (trained with the training set, number of neighbors = 5).

| **Cohort** | **Imputed features** |
| --- | --- |
| BioFINDER-1 | plasma P-tau231 plasma NTA plasma GFAP plasma NfL |
| ADNI | *APOE* E2 |
| UCSF-ADRC-ADRC | Delayed 10-word recall |
| A4 | Delayed 10-word recall  *APOE* E2  *APOE* E4 |
| OASIS | Delayed 10-word recall |
